# Whole genome mutational analysis for tumor-informed ctDNA based MRD surveillance, treatment monitoring and biological characterization of urothelial carcinoma

**DOI:** 10.1101/2023.07.13.23292590

**Authors:** Iver Nordentoft, Sia Viborg Lindskrog, Karin Birkenkamp-Demtröder, Santiago Gonzalez, Maja Kuzman, Jurica Levatic, Dunja Glavas, Ryan Ptashkin, James Smadbeck, Danielle Afterman, Tomer Lauterman, Yarin Cohen, Zohar Donenhirsh, Iman Tavassoly, Ury Alon, Amanda Frydendahl Boll Johansen, Mads Heilskov Rasmussen, Claus Lindbjerg Andersen, Paz Polak, Asaf Zviran, Boris Oklander, Mads Agerbæk, Jørgen Bjerggaard Jensen, Lars Dyrskjøt

**Affiliations:** Department of Molecular Medicine, Aarhus University Hospital, Aarhus, Denmark; Department of Clinical Medicine, Aarhus University, Aarhus, Denmark; C2i Genomics. INC, New York, NY, USA; C2i Genomics. LTD, Haifa, Israel; Department of Oncology, Aarhus University Hospital, Aarhus, Denmark; Department of Urology, Aarhus University Hospital, Aarhus, Denmark

**Author notes:** authors contributed equally (Iver Nordentoft, Sia Viborg Lindskrog).

## Abstract

Circulating tumor DNA (ctDNA) can be used for sensitive detection of minimal residual disease (MRD). However, the probability of detecting ctDNA at low tumor burden is limited by the number of mutations analyzed and available plasma volume. Here we applied a tumor-informed whole genome sequencing (WGS) approach for ctDNA-based MRD detection (91% sensitivity, 92% specificity) and treatment response evaluation in 916 longitudinally collected plasma samples from 112 patients with localized muscle-invasive bladder cancer. We show that WGS-based ctDNA detection is prognostic of patient outcomes with a median lead time of 131 days over radiographic imaging. We performed genomic characterization of post-treatment plasma samples with a high ctDNA level and observed acquisition of the platinum therapy-associated mutational signatures and copy number variations not present in the primary tumors. Our results support the use of WGS for ultra-sensitive ctDNA detection and highlight the additional possibility for plasma-based tracking of tumor evolution.

**Statement of significance:** Our study supports the clinical potential of using a WGS-based strategy for sensitive ctDNA detection in patients with MIBC. Thus, WGS-based ctDNA detection constitutes a promising option for clinical use due to low requirements for plasma input and the ease of performing WGS, eliminating the need for personalized assay design.

## Introduction

Tumor cells release cell-free DNA (cfDNA) with tumor specific molecular alterations into circulation (circulating tumor DNA; ctDNA) mainly by cell death^1^. ctDNA is cleared from circulation through nuclease digestion, renal clearance, or uptake by the liver and spleen^2–5^. Furthermore, the ctDNA half-life is between 15 min and 2 hours^6^, making it possible to use ctDNA for real-time tracking of tumor burden following surgery and through oncological treatment. Recent studies have shown that ctDNA is a powerful biomarker for detection of minimal residual disease (MRD) and recurrence in multiple cancer types^7,8^ – including bladder cancer (BC)^9^. Globally, more than 570,000 patients are diagnosed with BC each year^10^. Curative intended radical cystectomy (RC) preceded by neoadjuvant chemotherapy (NAC) is the standard of care for localized muscle-invasive bladder cancer (MIBC). However, nearly half of patients will experience metastatic relapse mainly within the first 2 years after surgery, when considering all stages^11^. Therefore, reliable diagnostic and prognostic biomarkers with high sensitivity and specificity are needed to improve detection of MRD for earlier initiation of oncological treatment and potentially improve patient outcomes. Previous ctDNA studies in patients with BC have demonstrated that ctDNA can be detected on average 3 months prior to metastatic relapse detected by radiographic imaging^9^. In addition, ctDNA has been correlated to chemo- and immunotherapy response^9,12,13^. The low tumor fraction typically observed post-surgery limits the probability of detecting ctDNA-based MRD. Tumor-informed ctDNA detection approaches have the highest sensitivity and specificity, but are also usually more labor intensive compared to non-tumor informed detection methods. Furthermore, the probability of detecting ctDNA is limited by the number of mutations analyzed, the available plasma volume and the depth of sequencing^14^. Recent studies have demonstrated the feasibility of using tumor-informed whole genome sequencing (WGS)-based analysis with a joint utilization of mutations and copy number alterations for ultra-sensitive ctDNA detection^15,16^. In addition, WGS-based analysis of ctDNA also provides the opportunity for genomic characterization of metastatic tumor biology to monitor tumor evolution and genomic changes acquired during oncological treatment^17^.

Here, we implemented and applied a WGS approach for sensitive ctDNA-based MRD detection and treatment response prediction in 112 patients with localized MIBC. Furthermore, we performed a *de novo* genomic characterization of plasma samples with a ctDNA level above 10% (n=15) to explore how WGS of plasma samples can be used to track tumor evolution.

## Results

### Patient characteristics and WGS data generation

A total of 112 patients with localized MIBC treated with NAC before RC were prospectively enrolled for liquid biopsy analysis between 2014 and 2021 at Aarhus University Hospital, Denmark (**Supplementary Table S1**). Plasma samples for ctDNA analysis were procured before, during and after NAC, and at scheduled control visits after RC (n = 916; **Fig. 1a**). RC was performed for 110 of the patients and these patients had a median follow-up time of 53.6 months after RC. We observed a recurrence rate of 24% (26/110), pathologic downstaging to a non-invasive stage (≤pTa,CIS,N0) for 61% (67/110) and complete pathologic response (pCR, pT0/pTIS) for 58% (64/110). WGS of tumor- and matched PBMC DNA was performed at a mean genome coverage of 59x and 33x for formalin fixed paraffin embedded (FFPE) and fresh frozen samples, respectively, and 31x for PBMC DNA. WGS of cfDNA from plasma (n=916) was performed at a mean genome coverage of 28x.

**Fig. 1:**
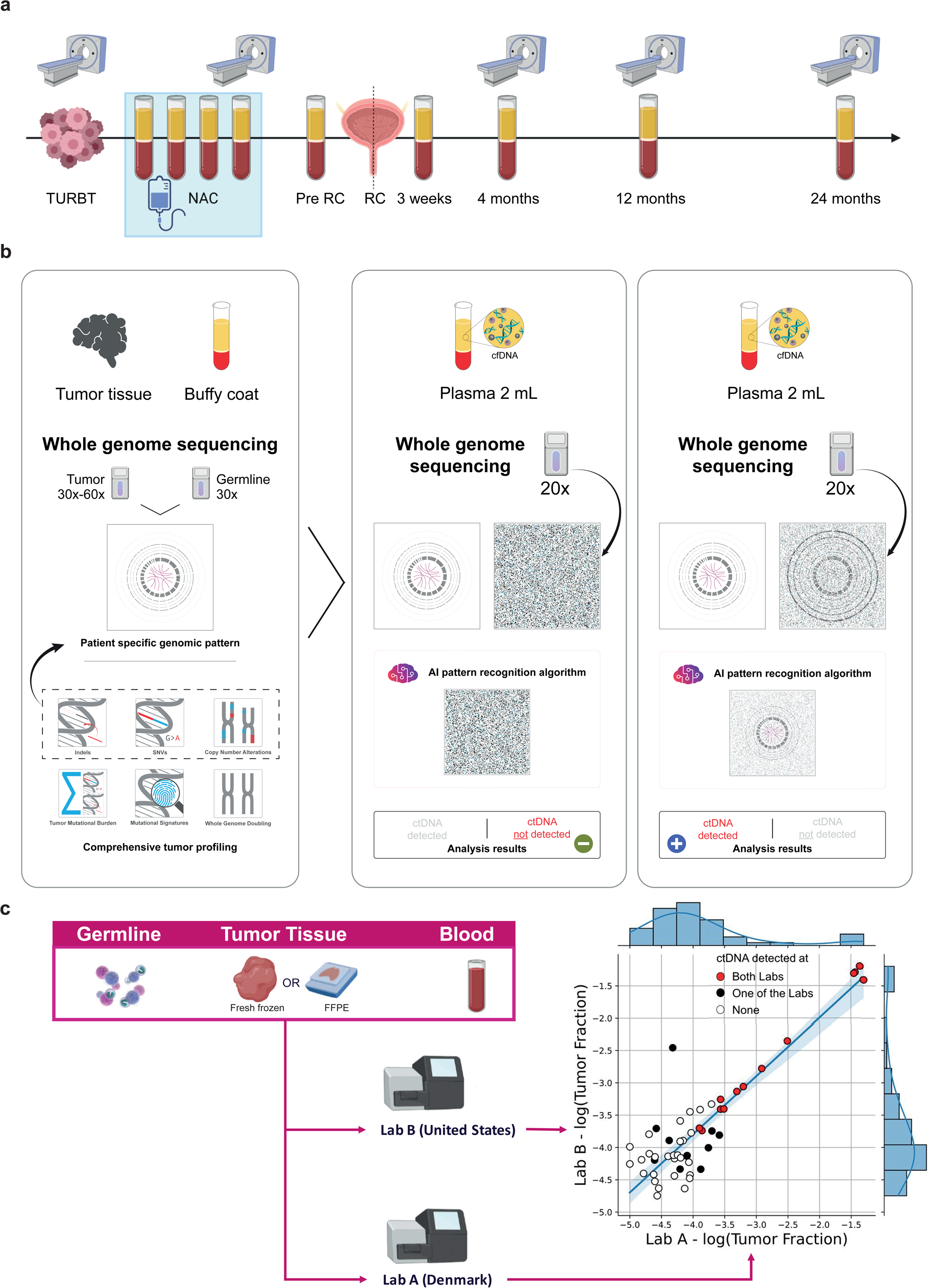
Study design and analysis scheme. **a**, The study design showing scheduled CT scanning (thorax-abdomen) and clinical sample collection. To exclude distant metastasis, full body PET/CT scans were performed after diagnosis. **b**, WGS approach for ctDNA detection. WGS of tumor/germline pairs were performed and patient-specific tumor genomic patterns were generated using signals from genome-wide alterations and AI-based error suppression (panel 1). The patient-specific tumor fingerprints were used for determining presence or absence of ctDNA in WGS data from plasma cfDNA (panel 2 and 3). **c**, Reproducibility of the method was established through independent processing of the tumor, germline and blood samples at two different laboratories in Denmark and The United States. The comparison included 52 blood samples from 18 patients. The coefficient of determination was R^2^=0.8 (all samples), and R^2^=0.99 (samples detected positive at both laboratories, red circles).

### ctDNA detection by tumor-informed WGS models

For ctDNA detection we developed patient-specific, tumor-informed WGS models by integrating genome-wide somatic alteration patterns coupled with advanced signal processing and AI-based error suppression (see materials and methods; **Fig. 1b**). The patient-specific models were applied to WGS data from plasma cfDNA for ctDNA detection. For initial validation of the robustness of the WGS ctDNA analysis pipeline, we analyzed technical replicates (tumor-, PBMC- and plasma DNA) performed independently in two different laboratories (USA, DK) using similar protocols and compared the ctDNA calls of 166 plasma samples from 18 patients (**Fig. 1c**). Of these, 52 samples had a tumor fraction above the detection threshold in one or both laboratory sites. A high correlation of estimated tumor fractions between laboratory sites was observed, with the coefficient of determination (R^2^) being 0.8 when including all samples and 0.99 when restricting to samples detected positive in both laboratory sites (red circles, **Fig. 1c**).

### WGS-based ctDNA detection for prognosis and metastatic relapse

The prognostic value of ctDNA was investigated using the 916 plasma samples collected during the disease courses of the 112 patients: i) at diagnosis prior to NAC (preNAC), ii) after NAC and before RC (preRC) and iii) during disease surveillance after RC (postRC; **Fig. 2**, (**Supplementary Fig. S1**). Detection of ctDNA was highly prognostic of patient outcomes: at diagnosis before NAC (Recurrence-free survival [RFS]: HR=7.7, 95%CI=2.3-26.3, *p*=0.0001; overall survival [OS]: HR=9.2, 95%CI=2.7-31.5, *p*=0.0001), at preRC (RFS: HR=3.4, 95%CI=1.5-7.8, *p*=0.0018; OS: HR=4, 95%CI=1.8-8.8, *p*=0.0003), and postRC (accumulated ctDNA status up to 1 year after RC; RFS: HR=23, 95%CI=7.9-67.1, *p*<0.0001; OS: HR=31.6, 95%CI=10.8-92.9, *p*<0.0001 (**Fig. 3, Supplementary Table S2**).

**Fig. 2:**
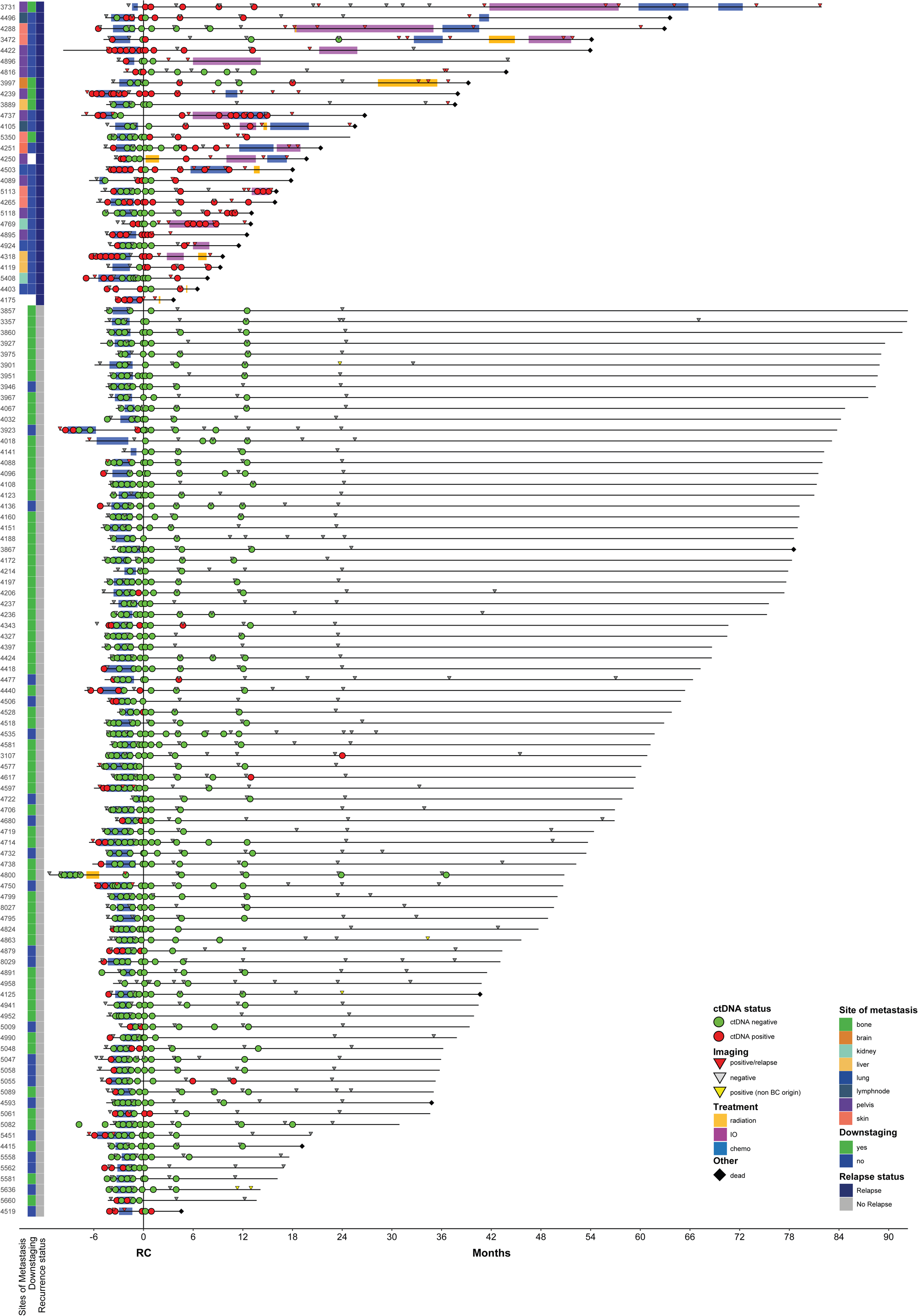
Longitudinal ctDNA results for all patients. Horizontal lines represent the disease courses of the patients, and circles represent ctDNA status. Treatment and imaging information are indicated for each patient (see color key). Patients are ordered by decreasing overall survival for patients with and without disease recurrence. Patients 4175 and 4250 were not able to undergo RC and time zero for these patients is the scheduled time for surgery.

**Fig. 3:**
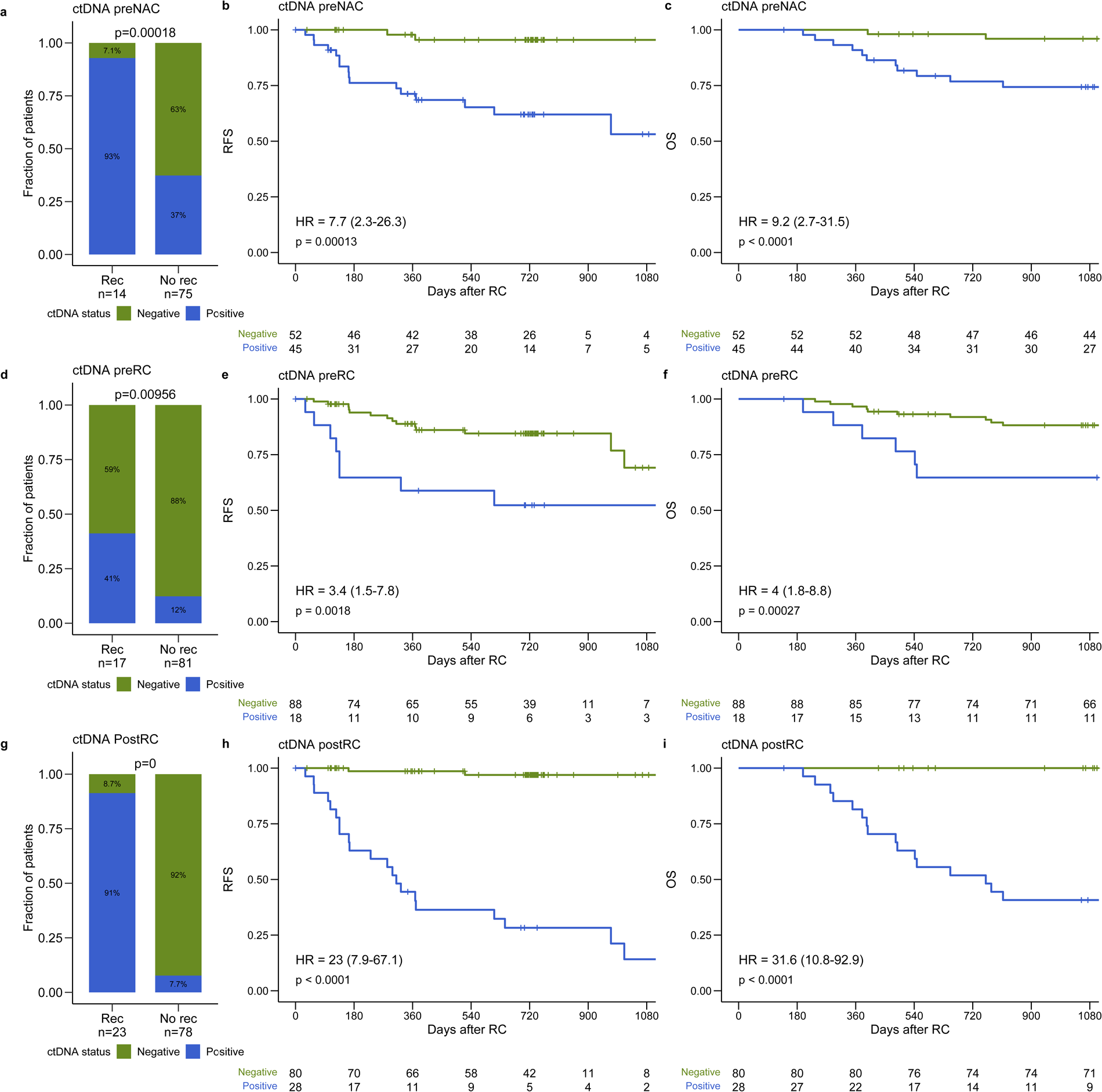
ctDNA detection for prognosis assessment. **a**, Association between plasma ctDNA status before NAC and recurrence status within one year after RC including only patients with at least two years of follow-up after RC for patients without recurrence. **b**, Kaplan-Meier survival analysis of RFS and plasma ctDNA status before NAC. **c**, Kaplan-Meier survival analysis of OS and plasma ctDNA status before NAC. **d**, Association between plasma ctDNA status before RC (after NAC) and recurrence status within one year after RC including only patients with at least two years of follow-up after RC for patients without recurrence. **e**, Kaplan-Meier survival analysis of RFS and plasma ctDNA status before RC. **f**, Kaplan-Meier survival analysis of OS and plasma ctDNA status before RC. **g**, Association between accumulated plasma ctDNA status up to the one year post RC visit and recurrence status within 18 months after the last plasma sample was analyzed for ctDNA. Only patients without recurrence with at least 18 months of follow-up after the last plasma sample were included. **h**, Kaplan-Meier survival analysis of RFS and accumulated plasma ctDNA status after RC. **i**, Kaplan-Meier survival analysis of OS and accumulated plasma ctDNA status after RC. Hazard ratios (HR) and associated 95% confidence intervals (CI) and p-values are displayed on each Kaplan-Meier plot (cox regression analysis). A significant statistical difference between ctDNA status and recurrence was determined using Fisher’s exact test.

The presence of ctDNA after RC, indicative of MRD, showed the highest correlation to outcome. Using accumulated ctDNA status up to one year after cystectomy and metastatic relapse within 18 months of the last plasma sample resulted in a sensitivity of 91% and a specificity of 92% (**Fig. 3g**). Using the same criteria, pathologic downstaging predicted metastatic relapse with a sensitivity of 83% and specificity of 77% (Data not shown). In 70% of patients with metastatic relapse (18/26), ctDNA was detected before clinical recurrence (radiographic imaging positive; **Fig. 2**). Overall, the median lead time of ctDNA detection was 131 days (−106 to 1156, *p*<0.00001) over radiographic imaging for patients with detectable ctDNA after RC (full follow-up included; **Fig. 4a**). In this study, plasma samples were longitudinally monitored after surgery for up to one year for non-recurrence patients. To assess if longer serial monitoring of ctDNA after RC would add an additional benefit, we calculated the cumulative incidence of ctDNA detection after RC (**Fig. 4b**). We observed that among patients with recurrence (n=26), the cumulative ctDNA detection was 85% at one year after RC compared to 6% for patients without recurrence (n=82, *p*<0.0001). Notably, almost all ctDNA-detected recurrences were found within the first 6 months after RC (95%, 21/22) (**Fig. 4b**).

**Fig. 4:**
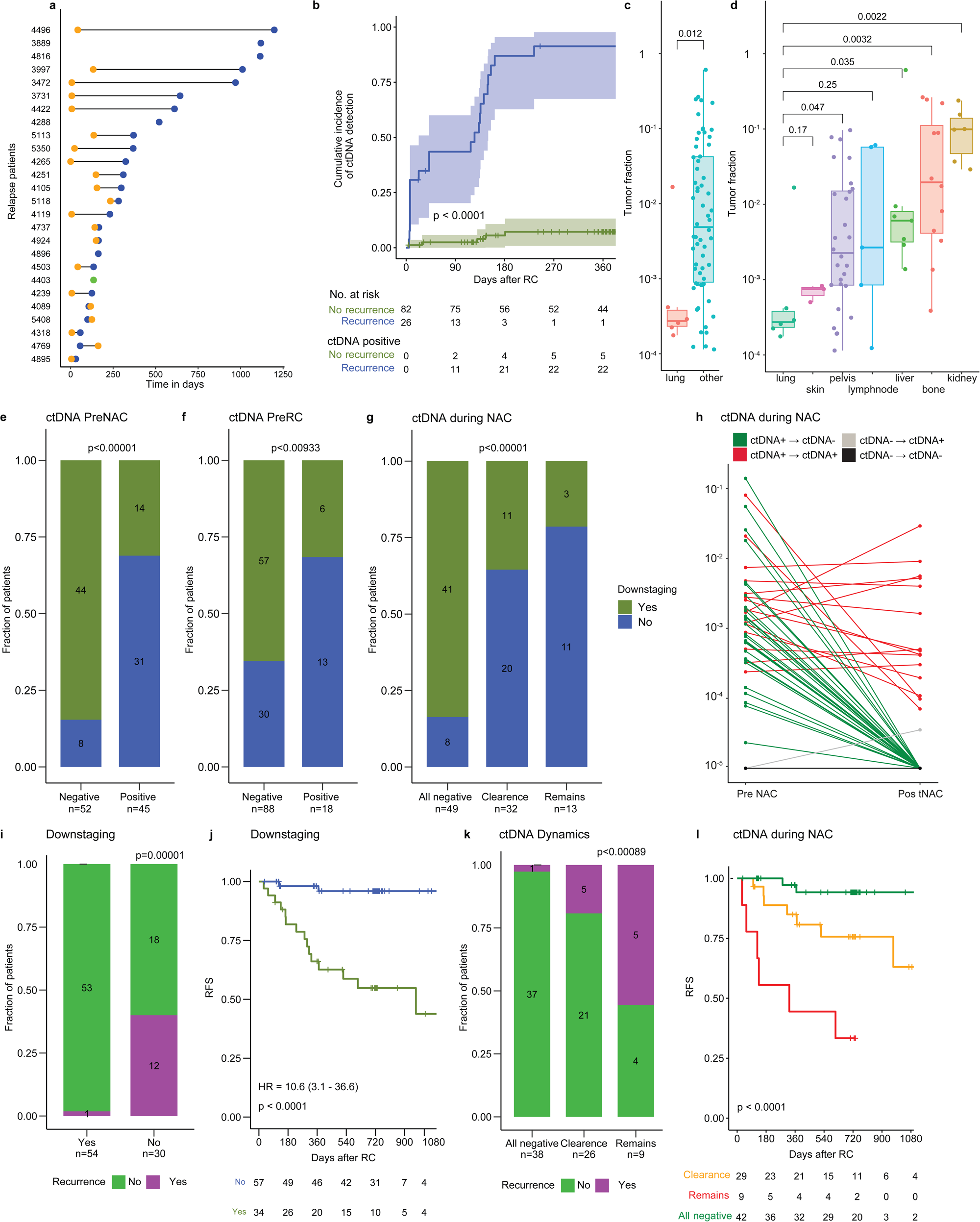
ctDNA measurements for detecting metastatic relapse and treatment response. **a,** Lead time in days between molecular recurrence (ctDNA positivity) and clinical recurrence (radiographic imaging positive). Statistical significance was calculated using paired Wilcoxon Rank test. **b,** Cumulative incidence of ctDNA detection after RC. **c,** Association between tumor fraction (ctDNA) and different metastatic sites. Only post-RC samples with a tumor fraction above zero were included. **d,** Association between tumor fraction (ctDNA) and lung metastases versus other metastatic sites. **e,** Association between ctDNA status before NAC and pathological downstaging. **f,** Association between ctDNA status before RC and pathological downstaging. **g,** Association between ctDNA clearance after NAC and pathological downstaging. **h,** Comparison of tumor fractions (ctDNA) at the pre- and post-NAC timepoints. Colors indicate whether ctDNA remained detectable (red), remained undetectable (black), were cleared during NAC (green), or appeared (grey). **i,** Association between pathological downstaging and recurrence status within one year after RC for patients who received minimum 3 series of NAC. **j,** Kaplan-Meier survival analysis of RFS and pathological downstaging for patients that have received minimum 3 cycles of NAC. **k,** Association between ctDNA clearance after NAC and recurrence status within one year after RC for patients who received minimum 3 series of NAC. **l,** Kaplan-Meier survival analysis of RFS and ctDNA clearance after NAC for patients who received minimum 3 series of NAC.

As the level of ctDNA detected after RC varied, we hypothesized that the tumor fraction could be related to the metastatic site. Interestingly, significantly lower tumor fractions were detected for patients with lung metastasis compared to all other metastatic sites (*p*=0.012, **Fig. 4c**) as well as compared to metastases from bone (*p*=0.003), kidney (*p*=0.002), liver (*p*=0.035) and pelvis (*p*=0.047) individually (**Fig. 4d**).

### ctDNA detection for evaluation of treatment response

ctDNA status preNAC and preRC was significantly associated with pathologic downstaging (*p*<0.00001 and *p*=0.009, respectively; **Fig. 4e,f**) and pCR (*p*<0.00001, *p*=0.038, data not shown). ctDNA dynamics during NAC, i.e. whether ctDNA remained detectable (remains), remained undetectable (all negative) or was cleared (clearance) was also a predictor of pathologic downstaging (*p*<0.00001, **Fig. 4g,h**). Pathologic downstaging was *per se* a strong predictor of metastatic relapse with a recurrence rate of 2% (1/54) and 40% (12/30) for patients with and without pathologic downstaging, respectively (**Fig. 4i,j**). While the pathologic evaluation of NAC response uses the local tumor response as a proxy of systemic response, ctDNA status preRC provides a measurement for local response and metastatic disease burden. Thus, we investigated ctDNA as a treatment response parameter for distant micrometastatic disease. The analysis was restricted to patients receiving ≥3 NAC cycles to ensure adequate treatment. Patients remaining ctDNA negative throughout NAC (all negative, n=38) had a very favorable outcome with only one patient experiencing metastatic recurrence (**Fig. 4k**). For patients with ctDNA clearance during NAC, 19% (5/26) of patients recurred. Finally, patients who remained ctDNA positive during NAC, had an unfavorable outcome with a recurrence rate of 55% (5/9). Plasma ctDNA dynamics was furthermore strongly associated with RFS, with a particularly poor outcome for patients where ctDNA remained detectable after NAC (**Fig. 4l**). The association between ctDNA detection during the patients’ disease courses, pathologic response to NAC and recurrence are shown using alluvial plots in **Supplementary Fig. S2**.

### Genomic characterization of primary tumors

WGS analysis of the primary tumor for 112 patients revealed a median of 23,851 single-nucleotide variants (SNVs) and 674 insertions and deletions (indels) per tumor, which is comparable to a previous WGS analysis of bladder tumors^18^. An overview of selected genomic alterations from known bladder driver genes^19^ is shown in **Fig. 5a**. *TP53*, *RB1*, *KMT2D*, *ARID1A* and *KDM6A* were the most frequently mutated genes and 55% of tumors had a *TERT* promoter mutation (**Fig. 5a**). In addition, 16 significantly mutated genes were identified using dndscv^20^, an algorithm that identifies potential cancer driver genes based on the dN/dS ratio with correction for trinucleotide mutation rates (**Supplementary Table S3**). Among the most significantly mutated genes, dndscv identified *TP53*, *RB1*, *KDM6A* and *ARID1A*, which highly overlapped with known BC drivers (highlighted with asterisks in **Fig. 5a**). Whole-genome doubling (WGD) was identified in 51% of tumors (57/112) by separating tumors based on ploidy and the level of heterozygosity (**Supplementary Fig. S3a**)^21,22^. Tumors with WGD had a significantly higher number of SNVs (*p*=0.013) and indels (*p*=0.0004) and more *TERT* mutations (*p*=0.02) when compared with near-diploid tumors. WGD has previously been associated with worse prognosis^23^; however, we observed no correlation to worse RFS or OS (**Supplementary Fig. S3b,c**).

**Fig. 5:**
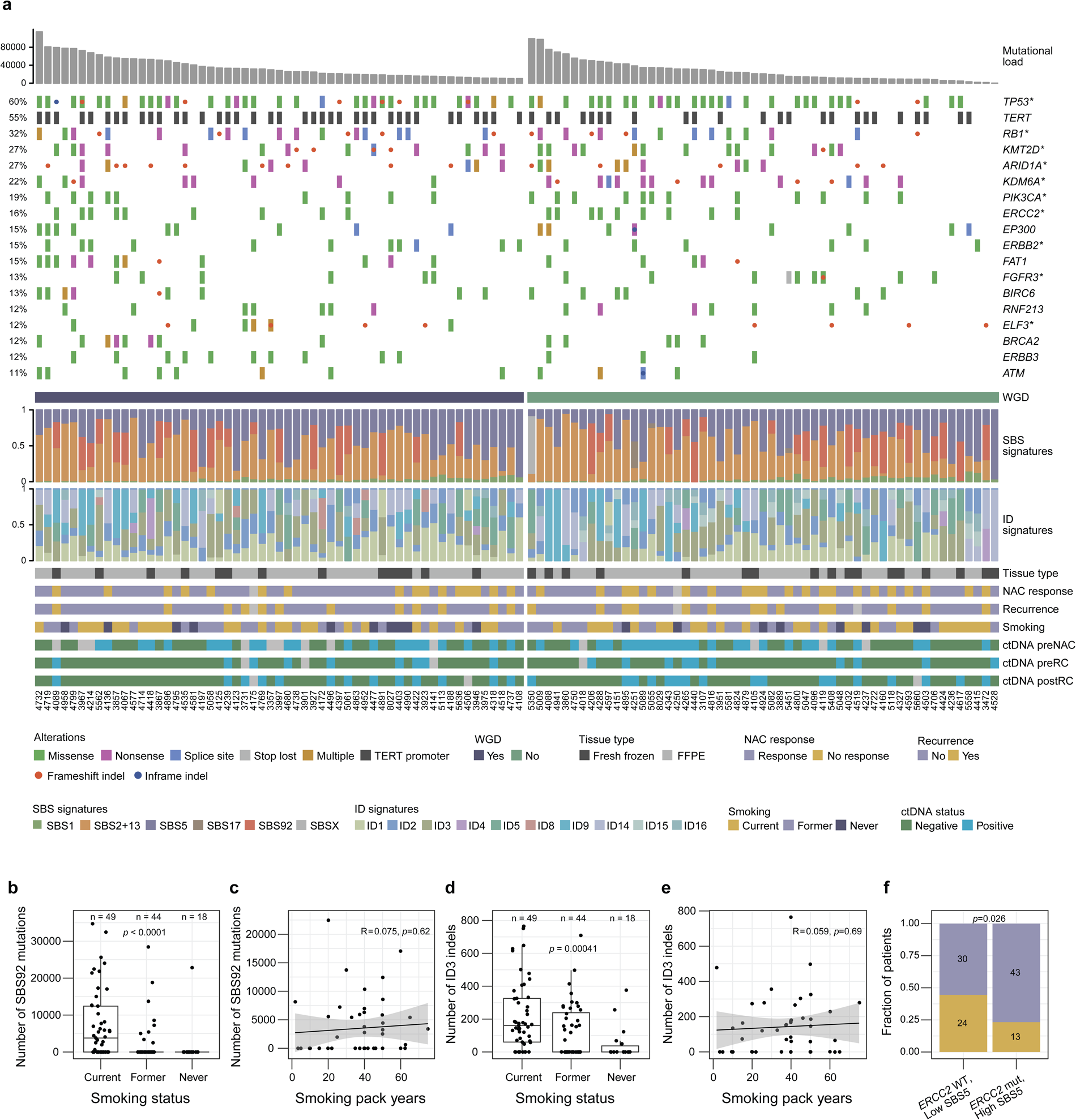
Genomic characterization of primary tumors. **a**, Genomic landscape of 112 primary tumors showing the mutational load including the number of single-nucleotide variants and indels, alterations in BC driver genes having a mutation frequency above 10% in the cohort, whole-genome doubling (WGD) status, contribution of single base substitution (SBS) signatures, contribution of small insertions and deletions (ID) signatures and clinical variables. Asterisks indicate genes significantly mutated in the cohort as defined by the dndscv algorithm. The tumor mutational load was defined as the total number of SNVs and indels. Tumors are divided by WGD status and sorted by decreased tumor mutational load. **b,** Number of mutations in the SBS92 context according to smoking status of the patients. **c,** Number of mutations in the SBS92 context versus the number of smoking pack years for current and former smokers. **d,** Number of small insertions and deletions in the ID3 context according to smoking status of the patients. **e,** Number of small insertions and deletions in the ID3 context versus the number of smoking pack years for current and former smokers. **f,** Association between pathological response to neoadjuvant chemotherapy (NAC) and the combination of *ERCC2* mutational status and SBS5 contribution (above or below the median SBS5 contribution).

*De novo* extraction of single-base substitutions (SBS) signatures revealed eight mutational signatures in the profiled tumor genomes, of which seven were decomposed to the known SBS1, SBS2, SBS5, SBS13, SBS17a, SBS17b, and SBS92. APOBEC-induced mutagenesis was identified as the primary contributor to the mutational landscape of the tumors with a median percentage of 42% of SNVs per patient being attributed to SBS2 or SBS13 mutational contexts (**Fig. 5a**). *De novo* extraction of small insertion-and-deletion (ID) signatures revealed 10 signatures of which seven were decomposed to the ID signatures previously observed in BC^26^ (ID1, ID2, ID3, ID4, ID5, ID8, ID9) and the remaining to the known ID14, ID15 and ID16 (**Fig. 5a**). ID1, ID3 and ID8 were identified as the primary contributors, each accounting for >15% of all observed indels. The mutational spectrum of tumors with WGD were enriched for mutations in the SBS1 (corrected *p*=0.014), SBS5 (corrected *p*=0.003), ID1 (corrected *p*=0.0003), ID2 (corrected *p*=0.003), ID6 (corrected *p*=0.04) and ID9 (corrected *p*=0.003) contexts when compared with near-diploid tumors.

The SBS92 signature was first identified in normal bladder tissue of smokers^24^ and has subsequently been attributed to tobacco smoking mutagenesis in BC using the PCAWG dataset^25^. Here, 45 tumors had SBS92-related mutations and we observed a significantly higher number of SBS92-related mutations in tumors from current smokers compared to tumors from former- and never smokers (*p*<0.0001; **Fig. 5b**). For current or former cigarette smokers, the number of mutations in the SBS92 context was not associated with the number of cigarette pack years (**Fig. 5c**). The SBS92 signature was also detected in plasma samples collected after RC for three patients with metastatic relapse (**Fig. 6a**). Furthermore, the ID3 signature has also previously been associated with tobacco smoking^26^. Here, 66 tumors had ID3-related indels and we observed a significantly higher number of ID3-related indels in tumors from current smokers compared to tumors from former- and never smokers (*p*=0.0004; **Fig. 5d**). Again, the number of indels in the ID3 context was not associated with the number of cigarette pack years (**Fig. 5e**). For current smokers, 90% (44/49) showed either SBS92- or ID3-related alterations and 51% (25/49) showed contribution from both signatures (**Supplementary Fig. S3d**). In comparison, only 28% (5/18) of never smokers showed contribution from either SBS92 or ID3, highlighting the divergent mutational landscape of tumors from smoking and non-smoking patients. No association between smoking status and number of SNVs or indels was observed (**Supplementary Fig. S3e,f**).

**Fig. 6:**
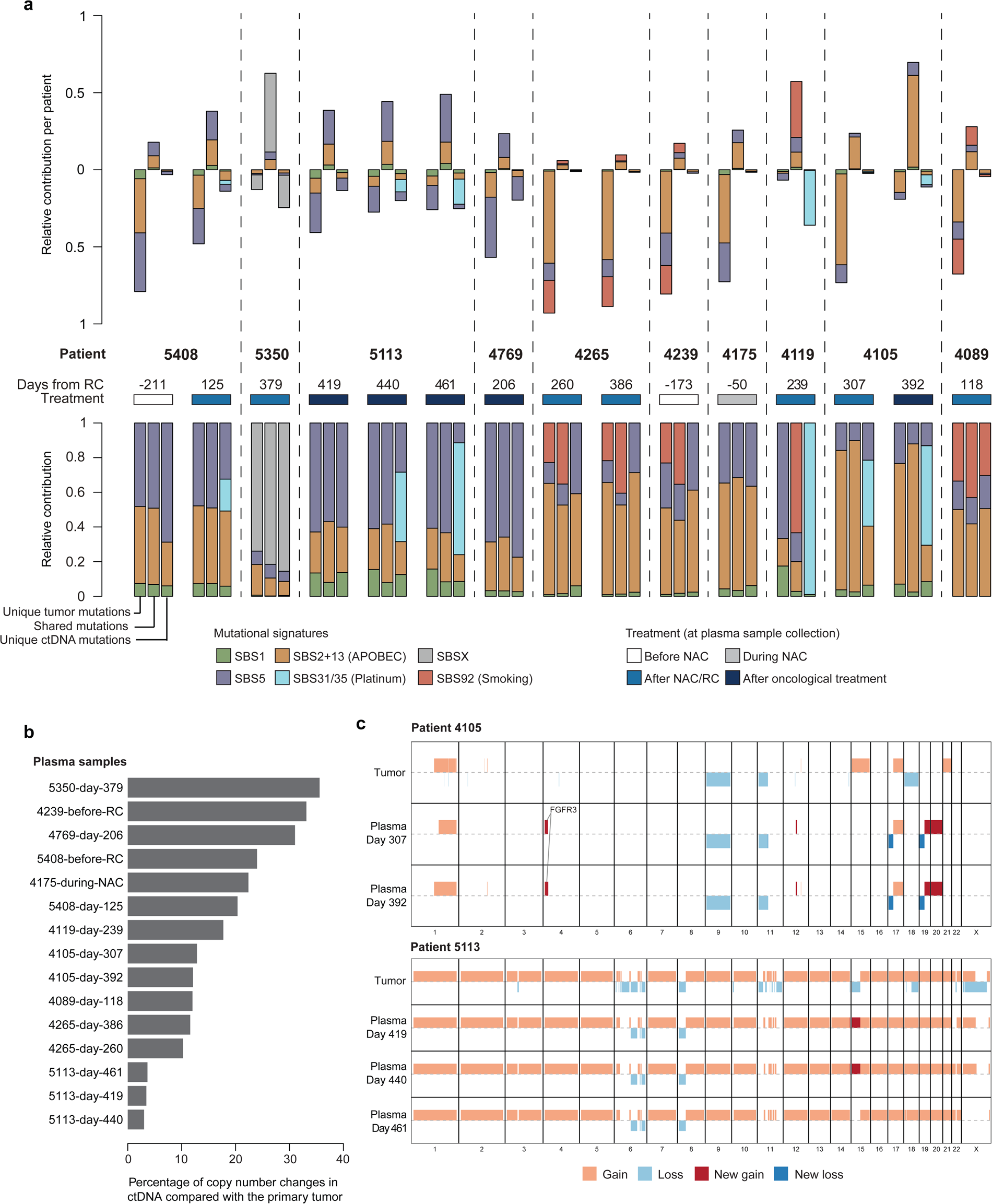
Genomic characterization of plasma samples with a high tumor fraction. **a**, Relative contribution of mutational signatures in primary tumors and plasma samples with a tumor fraction above 10%. Each patient is represented by three bars corresponding to the contribution of signatures for mutations present only in the primary tumor (left), mutations present both in primary tumor and ctDNA (middle) and mutations present exclusively in the ctDNA (right). The top panel indicates the fraction of mutations that are shared or unique to either tumor or plasma for each patient whereas the lower panel is normalized by the total number of SNVs per sample. The day for plasma sample collection in relation to RC is shown along with the treatment status at sample collection. **b,** The percentage of genome-wide copy number changes in ctDNA compared with the initial primary tumor sample for all plasma samples with a tumor fraction above 10%. **c,** Examples of changes in copy numbers comparing primary tumor (top row) versus ctDNA (following rows) for patient 4105 and 5113.

Of interest, patient 5350 displayed a novel SBS signature characterized by T>A and T>C mutations (referred to as SBSX), which was not present in the existing databases^27,28^ and could not be reconstructed by known signatures (**Supplementary Fig. S4a,b**). The same mutations contributing to this context were identified in a plasma sample taken 379 days after RC as well, thus confirming the presence of the signature (**Supplementary Fig. S4c,d**). The tumor from patient 5350 was among the most mutated in the cohort (98,036 SNVs) and the novel signature contributed to 80% of the total mutational burden (**Fig. 5a**). A deeper inspection of SBSX showed a strong transcriptional strand bias and enrichment in intergenic regions (**Supplementary Fig. S4e**). Similar patterns are observed for transcription coupled nucleotide excision repair (TC-NER) processes that are involved in UV and tobacco damage repair and may indicate that TC-NER is associated with this signature^29,30^. It is noteworthy that patient 5350 was diagnosed with a primary neuroendocrine carcinoma, which accounts for <1% of all BC cases and is an aggressive tumor type associated with poor prognosis^31^, and was treated for esophageal cancer seven years before the bladder tumor.

We compared genomic variables to chemotherapy response evaluations. We found no significant association between response to NAC and number of SNVs, indels or WGD (**Supplementary Fig. S3g-i**). As previously observed^32^, we confirmed a significant association between SBS5 contribution and *ERCC2* mutations (*p*<0.0001; **Supplementary Fig. S3j**). Although neither the level of SBS5-related mutations nor *ERCC2* mutations were independently associated with response to NAC (**Supplementary Fig. S3k,l)**, combining *ERCC2* mutations and the level of SBS5-related mutations showed that 77% of patients with an *ERCC2* mutation and/or high SBS5 contribution (above median) showed pathological downstaging after NAC, whereas patients with *ERCC2* wildtype and/or low SBS5 contribution had a response rate of 56% (*p*=0.026; **Fig. 5f**).

### Tumor evolution resolved from WGS-based ctDNA analysis

WGS of cfDNA from plasma samples with high tumor fraction allows for characterization of tumor evolution and therapy-induced genomic changes^33,34^. In 38% of patients with metastatic relapse (10/26), the detected cfDNA tumor fraction was sufficient (>10%) to perform a post-treatment genomic characterization independently of their initial tumor biopsy. In total, we performed *de novo* somatic variant calling for 15 plasma samples from those 10 patients to explore the potential for plasma-based characterization of tumor evolution using ∼20x WGS. Six plasma samples from four patients (5408, 5113, 4119 and 4105; 40% of included plasma samples) showed contribution of the platinum therapy-associated mutational signatures SBS31 or SBS35 (**Fig. 6a**). Importantly, the two chemotherapy-induced mutational signatures were not observed in any of the primary tumors and were only observed when analyzing plasma-specific mutations. For all four patients, the SBS31/35 signatures were observed in plasma samples collected 239-571 days after initiation of NAC (plasma samples collected earlier in the disease course of the patients were not characterized due to too low tumor fraction). None of the four patients had a pathological response to NAC; however, the presence of chemotherapy-induced mutational signatures in the plasma samples may indicate clonal expansions after the gemcitabine/cisplatin treatment.

The sequencing depth of ∼28x is a limitation for studying individual sites and the driver mutations observed in post-treatment plasma samples were mostly identical to the mutations detected in the primary tumors. New potential driver mutations were only detected for patient 5350, where mutations in *ASXL2* and *NFE2L2* were observed in the plasma sample collected 379 days after RC (tumor fraction of 50%). *ASXL2* is involved in chromatin remodeling and *NFE2L2* is a transcription factor involved in the regulation of oxidative stress and inflammatory responses, and mutations in these genes have previously been observed in BC^35,36^.

Remarkably, we observed an evolution of the CNVs detected in post-treatment plasma samples compared to the primary tumor for all 10 patients, suggesting that the metastatic lesion differs from the sequenced part of the primary tumor (**Fig. 6b**). In the two analyzed plasma samples collected after RC for patient 4105 (one of the patients having chemotherapy-induced mutational signatures), we observed a focal amplification on chromosome 4 affecting the primary tumor driver variant *FGFR3* p.S249C, and an increase in copy number on chromosomes 19q and 20 not detected in the primary tumor (**Fig. 6c**). In the plasma sample collected during NAC for patient 5408, newly acquired copy number gains on chromosomes 17, 19p and 22 were observed (**Supplementary Fig. S5**). Opposite to those large genomic changes, the three plasma samples collected after RC for patient 5113 were all highly representative of the primary tumor without minor recurrent acquired variations, indicating limited evolution after treatment (**Fig. 6c**).

## Discussion

We applied a WGS approach to monitor ctDNA for sensitive MRD detection in patients with localized MIBC, and documented the prognostic role of ctDNA at diagnosis, before RC and during surveillance. Our findings underline a role for ctDNA in guiding treatment decisions in BC in line with previous findings from other patient cohorts^9,12,13,37,38^. An important aspect of this WGS approach is its high sensitivity and specificity, which is comparable to other established tumor-informed tests applied in this setting, matched the ease of performing WGS without any need for designing personalized assays. WGS-based ctDNA analysis facilitates local sample processing and a rapid generation of test results and may thus drive the ctDNA field forward faster and pave the way for novel trials designs requiring immediate test results. Prospective ctDNA WGS analysis for MRD detection seems very promising with low false negative rates, and may soon be implemented for informed selection of patients for adjuvant treatments. In line with this, ongoing clinical trials will demonstrate if early ctDNA-guided treatment in the adjuvant setting is beneficial compared to treatment initiation upon detection of metastatic relapse based on radiographic imaging^39,40^. In addition, this study also supports the rationale behind investigating the benefit of NAC administration for ctDNA negative patients, and whether bladder sparing approaches can be applied based on ctDNA testing.

We identified significantly lower tumor fractions for patients with lung metastasis compared to other metastatic sites, indicating a correlation between extent of detectable ctDNA and location of metastasis. These findings are in accordance with a recent study on metastatic colorectal cancer showing that patients with lung-only and peritoneum-only metastatic disease had significantly lower levels of ctDNA compared to other metastatic sites^41^, indicating a decreased detection sensitivity. Early detection of MRD using WGS approaches may result in improved detection in BC patients with lung metastasis.

When exploring mutational signatures present in primary tumor biopsies, we identified the contribution of SBS92 to be associated with the smoking status of patients as previously observed in malignant and non-malignant bladder tissue^24,25^. SBS92 was recently identified in non-small cell lung cancer using deep WES (median of 413x)^42^; however, SBS92 has not previously been identified in exome-sequenced bladder tumors as the signature is mostly located in intergenic regions^25^. This highlights the importance of using WGS data to unravel the full genomic landscape of BC genomes. Patient 5350 displayed a novel signature, SBSX, which could not be reconstructed by known public databases of signatures. Although the signature was only observed in a single patient, we found the same signature in a post-treatment plasma sample as well, thus confirming that the signature is related to biology rather than being technically introduced. The identification of a novel signature contributing to 80% of the total mutational burden for patient 5330 claims the need for an exploration of whether the signature is present in larger patient cohorts.

Our WGS strategy for ctDNA-based MRD detection allows for direct genomic characterization of plasma cfDNA independently of the initial tumor biopsy. With increased sequencing depth and accompanying deep targeted sequencing of detected variants, this approach holds potential for increasing our understanding of tumor evolution and treatment resistance mechanisms. First, metastatic tumor biology can be fully unraveled. It has been shown that ctDNA contains multiple subclones and that synchronous metastatic tissue biopsies only comprise a small fraction of the total ctDNA^17^. Analysis of plasma samples can thereby overcome tissue sampling bias leading to clonal illusion and underestimated heterogeneity to recapitulate the complete metastatic disease burden of the patients^43^. Second, tumor evolution and therapy-induced shifts in selection pressure can be longitudinally tracked. Tumor biology evolves over time and during treatment pressure, underlining the importance of having a real-time snapshot of the current tumor biology when making treatment decisions in the metastatic setting. Analysis of plasma samples during treatment could thereby serve as a minimally-invasive measure of treatment response (ctDNA clearance) and could, potentially, provide additional information of possible oncological treatment options.

Here, we observed a contribution of the platinum therapy-associated mutational signatures SBS31 or SBS35 in six post-treatment plasma samples from four patients, indicating cisplatin mutagenesis and clonal expansions after NAC. Furthermore, in the post-treatment plasma samples for patient 4105, we observed an acquired focal amplification of the *FGFR3* gene on chromosome 4, indicating that Erdafitinib, a pan-FGFR inhibitor approved as second-line treatment for patients with advanced BC^44^, could be a potential treatment option for this patient. These observations highlight how post-treatment tumor characterization using cfDNA provides information on the genomic changes acquired since the initial tumor biopsy, which could improve identification of therapeutic targets and provide information on possible treatment resistance at an early time point. An increased sequencing depth is, however, required to improve the genomic characterization and study clinically actionable gene mutations. The sequencing depth of ∼28x is a limitation for studying individual sites, and acquisition of new driver mutations was only observed for patient 5350 (tumor fraction of 50%) in our current analysis of mutational driver events in post-treatment plasma samples. Increased sequencing depth may also allow for biological characterization of plasma samples with a tumor fraction below the currently applied cut-off of 10%. Furthermore, to build on our findings presented here, WGS of metastatic tissue biopsies could be performed to study whether ctDNA profiling provides a more complete genomic characterization of all metastatic tumor sites compared with analyzing a single metastatic lesion. A previous study performed deep WGS of plasma samples (187x) with high ctDNA fractions (17-82%) and synchronous metastatic tissue biopsies (98x) from patients with advanced prostate cancer^17^. Overall, the study demonstrated how deep WGS of plasma samples is a superior approach to achieve high resolution of treatment-associated dynamics and resistance mechanisms compared to analyzing metastatic tissue biopsies.

Our study highlights that WGS-based analysis of cfDNA allows ultra-sensitive ctDNA detection in patients with MIBC. ctDNA testing holds huge potential to change the current clinical management of patients towards more individualized treatment. Furthermore, we showed that the WGS-based ctDNA detection approach provides the opportunity to perform *de novo* characterization of genomic changes acquired post-treatment, which is not possible with bespoke ctDNA detection methods. Plasma-based tracking of tumor evolution may ultimately open opportunities to refine precision oncology.

## Materials and methods

### Patients and clinical follow-up

In total, 112 patients diagnosed with MIBC treated with NAC and RC were enrolled between 2013 and 2021 at Aarhus University Hospital in Denmark. Detailed follow-up data were available for all patients with a median follow-up of 1841 days after RC (range: 139-2810) for patients without clinical recurrence. Recurrence data were obtained from computed tomography scans (CT-scans, PET/CT scans) and/or pathology reports and survival data were obtained from the nationwide civil registry. pCR to NAC was defined as pT0/pTIS/N0 and non-invasive downstaging after NAC was defined as pT0/pTIS/Ta/N0. All patients provided informed written consent, and the study was approved by The National Committee on Health Research Ethics (#1302183 and #1706291). Study data were collected and managed using REDCap hosted at Aarhus University^45,46^.

### Biological samples

Tumor samples were procured from transurethral resection of the bladder (TURBT) at the time of diagnosis (n=112). DNA was extracted from sections of Tissue-Tek O.C.T Compound embedded tissue or punches of formalin-fixed paraffin embedded tissue (FFPE) using Puregene DNA purification kit (Gentra Systems). Leukocyte DNA was extracted from the buffy coat from all patients using the QIAsymphony DSP DNA midi kit (QIAGEN). Cell-free DNA (cfDNA) was extracted from 2 mL of plasma using QIAamp Circulating Nucleic Acid Kit (QIAGEN, Hilden, Germany) and eluated in 60 µL buffer EB (QIAGEN). Blood samples were collected at scheduled clinical visits. The cfDNA percentile was estimated from TapeStation 4200 (Agilent) by size window: (100bp – 500bp) using the Cell-free DNA ScreenTape assay. High molecular fragments >500bp as a measure of genomic DNA contamination served as exclusion criteria if >30%. Total cfDNA concentration was measured using ddPCR assays for 2 stable regions on chromosome 16 and chromosome 3^47^. Automated Droplet Generator (Bio-Rad) were used for Droplet generation, and readout was performed on a QX200™ Droplet Reader (Bio-Rad).

### Whole Genome Sequencing

Libraries of tumor and matching germline DNA were prepared using the Twist Library preparation EF kit (Twist Bioscience) with an input of 200 ng DNA. The protocol utilizes enzymatic fragmentation; however, the fragmentation time was decreased to 6 minutes for the FFPE samples to account for degraded DNA. For buffy coat DNA and DNA from fresh frozen tumors, a 10 minutes fragmentation time was used. The protocol was optimized using the xGEN™ UDI-UMI Adapters (Integrated DNA Technologies) with 7 cycles of PCR post ligation. The UMI part was not sequenced as UMI correction is only beneficial for deep sequencing. Libraries of cfDNA from plasma were prepared using the KAPA HyperPrep kit (Roche) with the xGEN™ UDI-UMI Adapters (Integrated DNA Technologies) using cfDNA input equal to 1-2 mL of plasma and 7 cycles of PCR post ligation. A minimum input of 5 ng was used. All libraries were paired-end sequenced (2x150 bp) on the NovaSeq 6000 platform (illumina) using S4 flow cells. Prior to sequencing all runs were calibrated on a MiSeq Nano (2x150 bp) to obtain even coverage.

### Preprocessing of WGS data and quality control analysis

WGS reads for primary tumors, matched germline and plasma samples were demultiplexed using Illumina’s bcl2fastq to generate FASTQ files. The genetic concordance of FASTQ files from the same patient was confirmed using NGSCheckMate^48^. FASTQ files from all three sample types were trimmed with Skewer v0.2.2^49^ to remove paired-end adapter sequences. Both the untrimmed and trimmed FASTQ files were run through FastQC v0.11.9 to identify potential problems in sequencing. The trimmed FASTQ files were aligned to the reference genome (GRCh38) with BWA MEM v 0.7.17. The resulting aligned bam files were sorted with Samtools v1.14.

Each per-lane BAM file was marked for duplicate reads using GATK^50^ MarkDuplicatesSpark v.4.1.8.0 resulting in a duplicate-marked BAM that was passed for calculation and recalibration of the per-read base quality score using GATK BQSRPipelineSpark. Each recalibrated BAM file was indexed and re-sorted by read name using Samtools v1.11. GATK MarkDuplicatesSpark was used to merge all BAM files from the same sample. This process produced the final coordinate sorted BAM file for each sample.

Alignment quality control metrics were computed on the BAM file using Picard ( QualityScoreDistribution, MeanQualityByCycle, CollectBaseDistributionByCycle, CollectAlignmentSummaryMetrics, CollectInsertSizeMetrics, CollectGcBiasMetrics, CollectOxoGMetrics) and GATK (average coverage, percentage of mapped and duplicate reads). These metrics were used to identify potential problems in sequencing or preprocessing.

### Tumor/normal somatic mutation calling

Each tumor and matched germline BAM files were analyzed using GATK Mutect2 v4.2.4.1 and Strelka v2.9.10^51^ to identify putative somatic single nucleotide variants (SNVs). These SNVs were filtered using GATK FilterMutectCalls to retain PASS variants and remove variants corresponding to known single nucleotide polymorphism sites (dbSNP v138), as annotated by GATK VariantAnnotator. Only SNVs detected by both Mutect2 and Stelka were retained. Finally, a panel of sequenced donor plasma samples was used to remove artifactual SNV calls from the final SNV vcf file.

Somatic INDELs were called by SVaba v1.1.3 and Mutect2 and the final list of INDELs comprised those detected by both methods. In cases where INDELs overlapped with a non-exact match, the largest INDEL was selected.

Copy number variations (CNVs) in solid tumors were called using FACETS v0.6.2^52^. Base coverage and variant allele fraction (VAF) of heterozygous SNP positions of the tumor sample and paired germline sample were used as input to the method. The list of germline heterozygous SNP positions was calculated using BCFTools mpileup^53^ and a reference set of SNPs from the HapMap Project v3.3^54^. The output was processed to revert all subclonal CNV calls to the nearest clonal CNV call.

### Estimation of tumor fraction in solid tumor and cfDNA

Tumor fraction estimations in both solid tumor samples and plasma samples were estimated using the in-house method C2inform which constitutes the latest version of our previously described method^15^. The approach is based on genome-wide detection of somatic mutations found in the patient’s tumor sample, including SNVs, INDELs and CNVs, which are used to generate a patient-specific tumor signature. Additionally, the method is using a proprietary error-suppression model, which is based on WGS of cfDNA from 45 healthy individuals. The technique facilitates the detection of tumor presence in cfDNA and aids in estimating the tumor fraction.

Accumulated ctDNA status up to one year after RC was defined as any ctDNA positive plasma sample during that time period. The prognostic potential of ctDNA detection after RC was evaluated with a restricted follow-up time of 18 months after the last ctDNA analysis. Thus, recurrence was defined as detection of clinical recurrence from radiographic imaging and/or pathology reports in the time period of 18 months after the last plasma sample was analyzed. For non-relapsing patients, only patients with at least 18 months of follow-up after the last plasma sample were included.

### Genomic characterization of tumor samples

For the purpose of tumor characterization, a gradient boosting model was trained using individual mutation quality metrics and applied to remove low-quality mutations with features characteristic of FFPE-induced artifacts. The filtering method removed 29% of the excessive mutations in the FFPE samples without affecting fresh frozen samples (2.7% of mutations removed) and the mutational context similarity between FFPE and fresh frozen samples was increased by 5% (**Supplementary Fig. S6A-B**). After filtering, no significant difference in the number of SNVs and indels per tumor were observed between FFPE and fresh frozen samples (**Supplementary Fig. S6C-D**). VEP v107.0^55^ was applied to the final list of SNVs and INDELs to determine the affected gene and functional impact. We queried all non-synonymous variants of the cohort against databases of cancer driver genes (Intogen^19^ and cancer biomarkers CGI^56^) to identify the somatic mutations in genes of biological interest.

Similarly, genes affected by CNVs were classified as biologically relevant if present in a list of candidate cancer genes created by manual annotation of scientific publications^57,58^.

Sigprofiler v1.1.20 was used to extract SNV and INDEL signatures in a three-step workflow: (I) De-novo extraction of signatures. (II) Fitting of selected signatures from COSMIC v3.3^26^ and artifact signatures used for the detection of spurious deviations. (III) A final fitting using cancer-type exclusive signatures and the manual inclusion of select signatures determined from steps (I) and (II), that corresponds to the final set of signatures presented in the results.

Whole genome doubling (WGD) status was determined by separating samples based on ploidy and level of heterozygosity as previously described^21,22^, resulting in two separate clusters corresponding to near-diploid samples and samples with WGD.

### De novo detection of somatic alterations in plasma samples with high tumor fraction

Plasma samples with a tumor fraction above 10% were selected for de-novo calling of SNVs and CNVs (See: Biological characterization of tumor samples). SNV and INDEL results from the plasma samples were compared with results from the solid tumor samples to determine shared, tumor-unique, and plasma-unique alterations.

### Statistical analysis

Survival and cumulative incidence curves were compared using the Kaplan-Meier method (log rank tests). Hazard ratios (HR) and associated 95% confidence intervals (CI) were calculated using Cox regression analysis (R packages survminer v0.4.9 and survival v3.2.13). Kruskal Wallis test, Wilcoxon rank sum test, Fisher’s exact test and Pearson’s correlation coefficient were used to determine statistically significant associations. Analysis was performed using the R statistical environment (v4.1.2).

## Data availability

The WGS data generated during the study is available through controlled access from GenomeDK under accession number GDK000007 (https://genome.au.dk/library/GDK000007/).

## Acknowledgements

This work was funded by research grants to L.D. from C2i Genomics, The Danish Cancer Society, Novo Nordisk Foundation, Independent Research Fund Denmark, Aarhus University and Aarhus University Hospital. We would like to thank technical personnel at the Departments of Molecular Medicine, Urology and Oncology, Aarhus University Hospital, for sample handling and processing. Sample collection was supported by the Danish Cancer Biobank.

## Author contributions

L.D., A.Z., B.O., I.N. conception and design. L.D., A.Z., B.O., I.N., A.F.B.J. development of NGS methodology. I.N., A.F.B.J. NGS library generation and WGS sequencing coordination, Acquisition of data. K.B.D. selection of patients and acquisition of samples and clinical data and programming and managing of Redcap database. I.N., S.V.L., K.B.D., L.D., S.G., P.P., D.A. analysis and interpretation of data. I.N., S.V.L. S.G., P.P., L.D. wrote the manuscript, with contributions from K.B.D., M.K., J.L., D.G., R.P., J.S., D.A., T.L., Y.C., Z.D., I.T., U.A., A.F.B.J., M.H.R., C.L.A., A.Z., B.O., M.A., J.B.J.

## Competing interests

Lars Dyrskjøt has sponsored research agreements with C2i Genomics, Natera, AstraZeneca, Photocure, and Ferring and has an advisory/consulting role at Ferring, MSD and UroGen. Lars Dyrskjøt has received speaker honoraria from AstraZeneca, Pfizer and Roche and received travel support from MSD. Lars Dyrskjøt is board member at BioXpedia. Jørgen Bjerggaard Jensen is a member of Advisory Boards at Ferring, Roche, Cepheid, Urotech, Olympus, AMBU, Janssen, and Cystotech, is a speaker at medac, Olympus, Intuitive Surgery, Photocure ASA, and has research collaborations with medac, Photocure ASA, Roche, Ferring, Olympus, Intuitive Surgery, Astellas, Cepheid, Nucleix, Urotech, Pfizer, AstraZeneca, MeqNordic, Laborie, VingMed, AMBU and Cystotech.

Asaf Zviran is the co-founder of C2i Genomics. Boris Oklander is the co-founder and CTO of C2i Genomics. Santiago Gonzalez, Maja Kuzman, Jurica Levatic, Dunja Glavas, Ryan Ptashkin, James Smadbeck, Danielle Afterman, Tomer Lauterman, Yarin Cohen, Zohar Donenhirsh, Iman Tavassoly, Ury Alon are employees of C2i Genomics. Paz Polak is a former employee of C2i Genomics.

## Notes

### Competing Interest Statement

Lars Dyrskjoet has sponsored research agreements with C2i Genomics, Natera, AstraZeneca, Photocure, and Ferring and has an advisory/consulting role at Ferring, MSD and UroGen. Lars Dyrskjoet has received speaker honoraria from AstraZeneca, Pfizer and Roche and received travel support from MSD. Lars Dyrskjoet is board member at BioXpedia.
Joergen Bjerggaard Jensen is proctor for Intuitive Surgery, is a member of advisory board for Olympus Europe, Ambu, Cepheid, Janssen, and Ferring and has sponsored research agreements with Medac, Photocure ASA, Cepheid, Olympus, and Ferring.
Asaf Zviran is the co-founder and a member of the board of directors of C2i Genomics. Boris Oklander is the co-founder and CTO of C2i Genomics.
Santiago Gonzalez, Maja Kuzman, Jurica Levatic, Dunja Glavas, Ryan Ptashkin, James Smadbeck, Danielle Afterman, Tomer Lauterman, Yarin Cohen, Zohar Donenhirsh, Iman Tavassoly, Ury Alon are employees of C2i Genomics. Paz Polak is a former employee of C2i Genomics.

### Author Declarations

All patients provided informed written consent, and the study was approved by The National Committee on Health Research Ethics (#1302183). Study data were collected and managed using REDCap hosted at Aarhus University

### Summary of Updates

The manuscript has been updated based on previous reviewer feedback

