## Supplementary figure S1-S6 and supplementary table S1-S3 for "Whole genome mutational analysis for tumor-informed ctDNA based MRD surveillance, treatment monitoring and biological characterization of urothelial carcinoma"

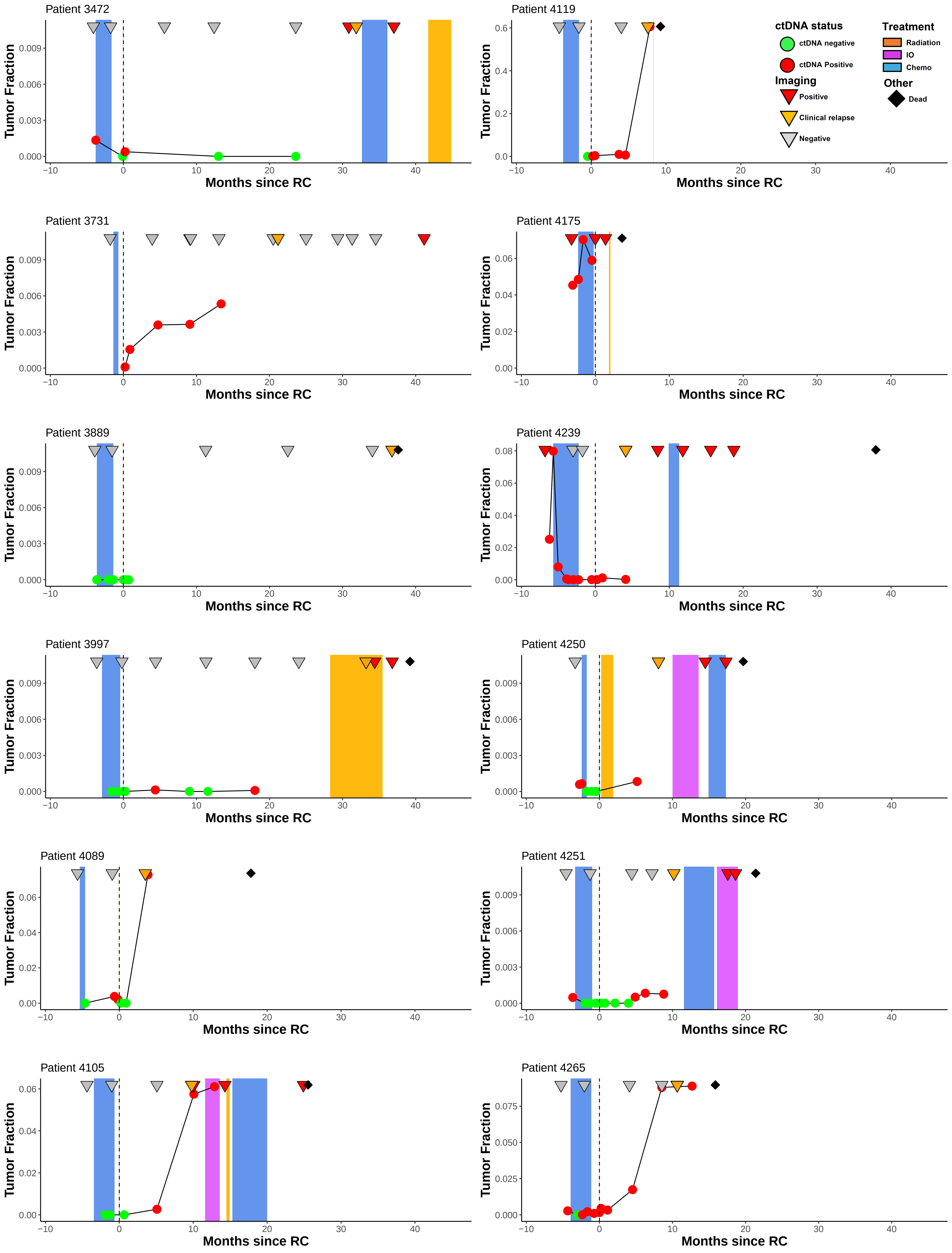
Patient 3472
Patient 4119
Patient 3731
Patient 4175
Patient 3889
Patient 4239
Patient 3997
Patient 4250
Patient 4089
Patient 4251
Patient 4105
Patient 4265
Tumor Fraction
Months since RC
ctDNA status
ctDNA negative
ctDNA Positive
Imaging
Positive
Clinical relapse
Negative
Treatment
Radiation
IO
Chemo
Other
Dead

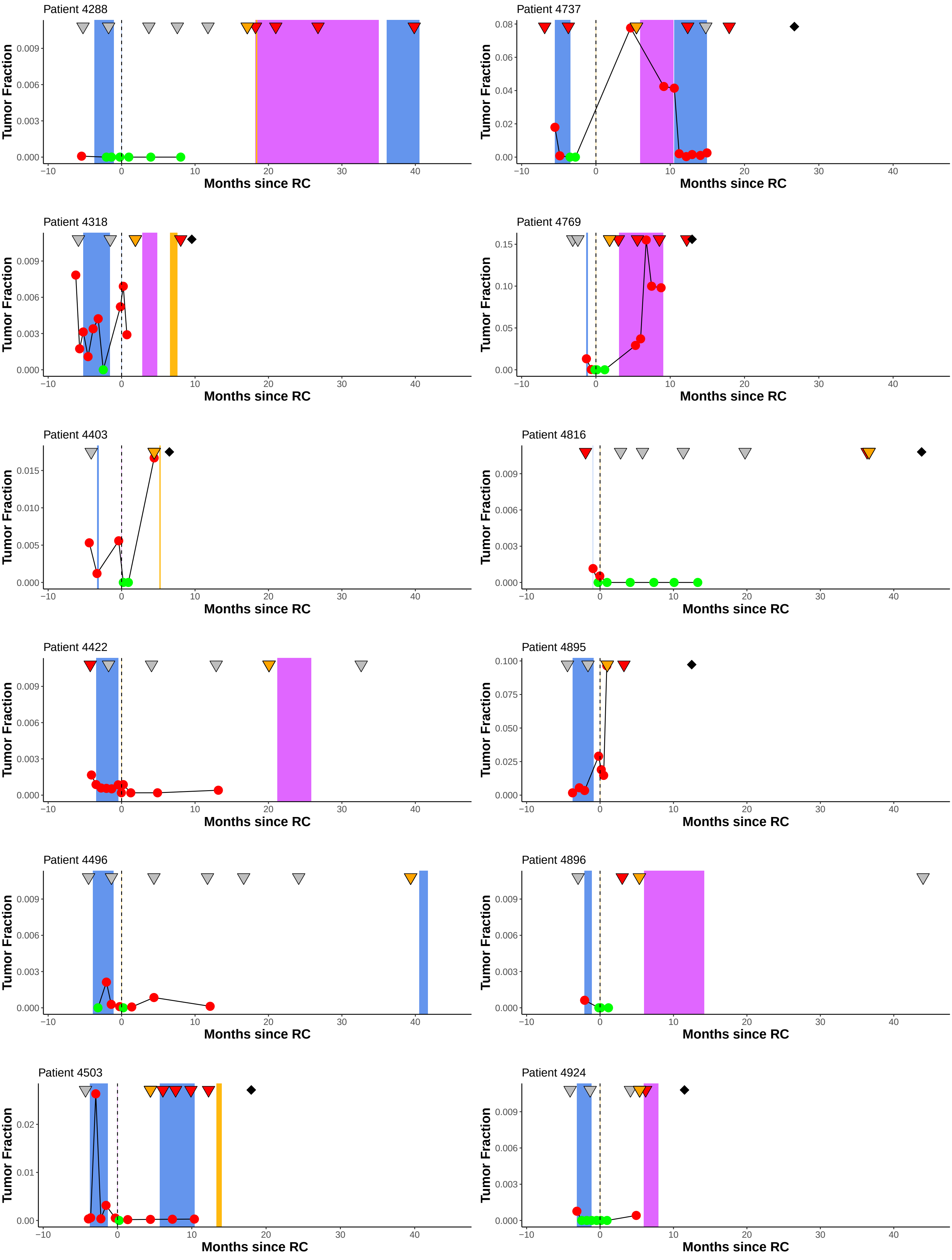
Patient 4288
Patient 4737
Patient 4318
Patient 4769
Patient 4403
Patient 4816
Patient 4422
Patient 4895
Patient 4496
Patient 4896
Patient 4503
Patient 4924
Tumor Fraction
Months since RC

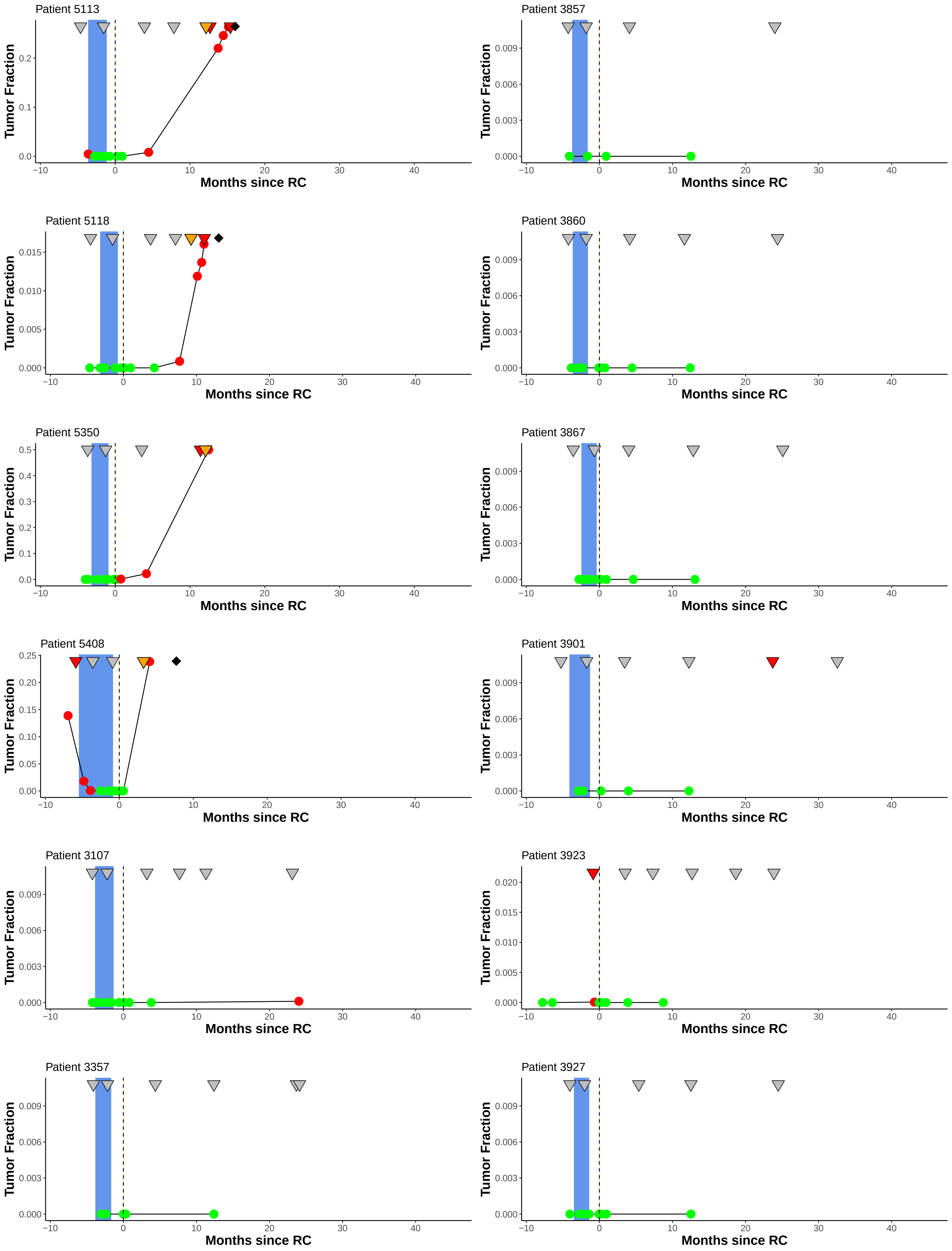

Patient 5113
Patient 3857
Patient 5118
Patient 3860
Patient 5350
Patient 3867
Patient 5408
Patient 3901
Patient 3107
Patient 3923
Patient 3357
Patient 3927
Tumor Fraction
Months since RC

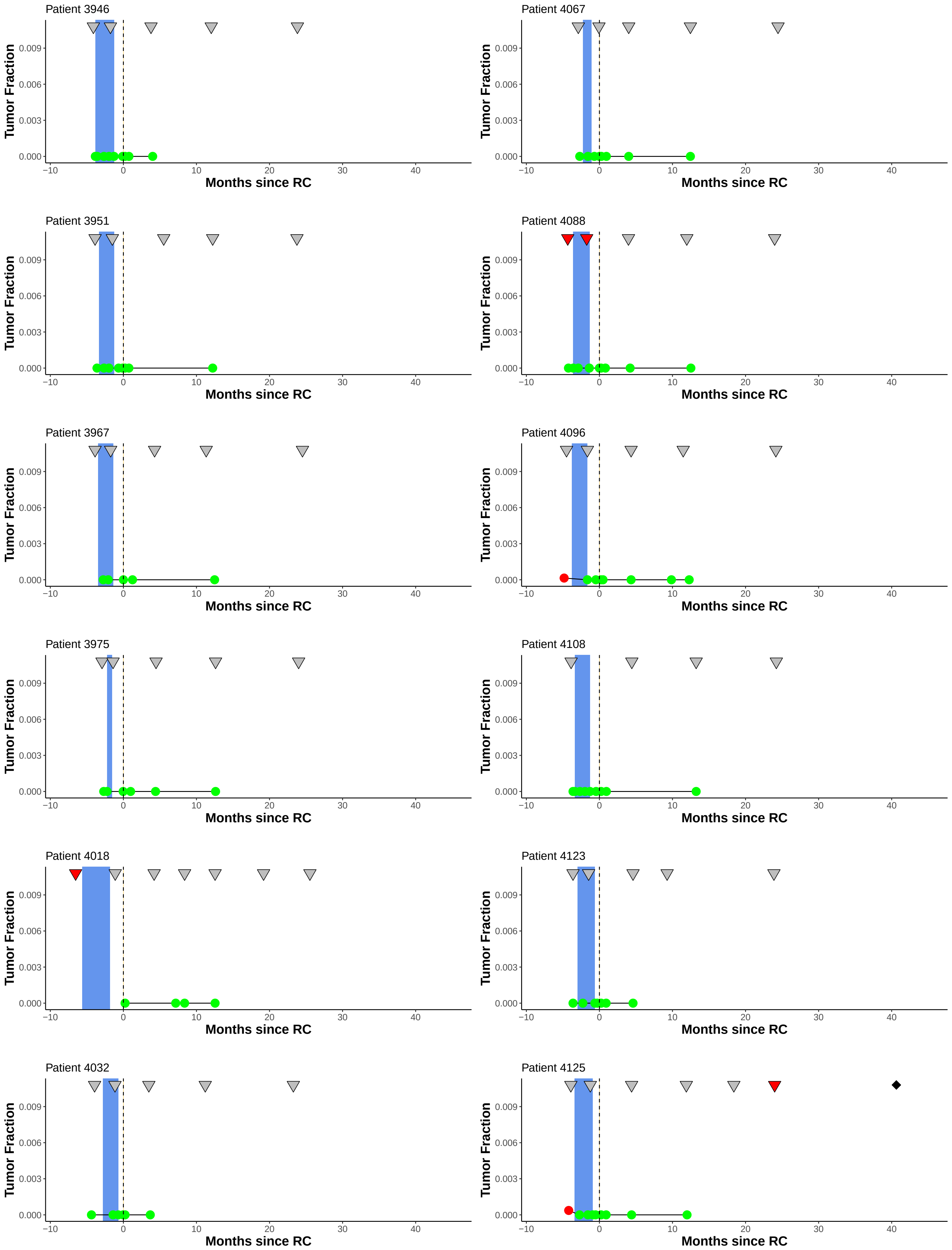

Patient 3946
Patient 4067
Patient 3951
Patient 4088
Patient 3967
Patient 4096
Patient 3975
Patient 4108
Patient 4018
Patient 4123
Patient 4032
Patient 4125
Tumor Fraction
0.000
0.003
0.006
0.009
Months since RC
−10
0
10
20
30
40

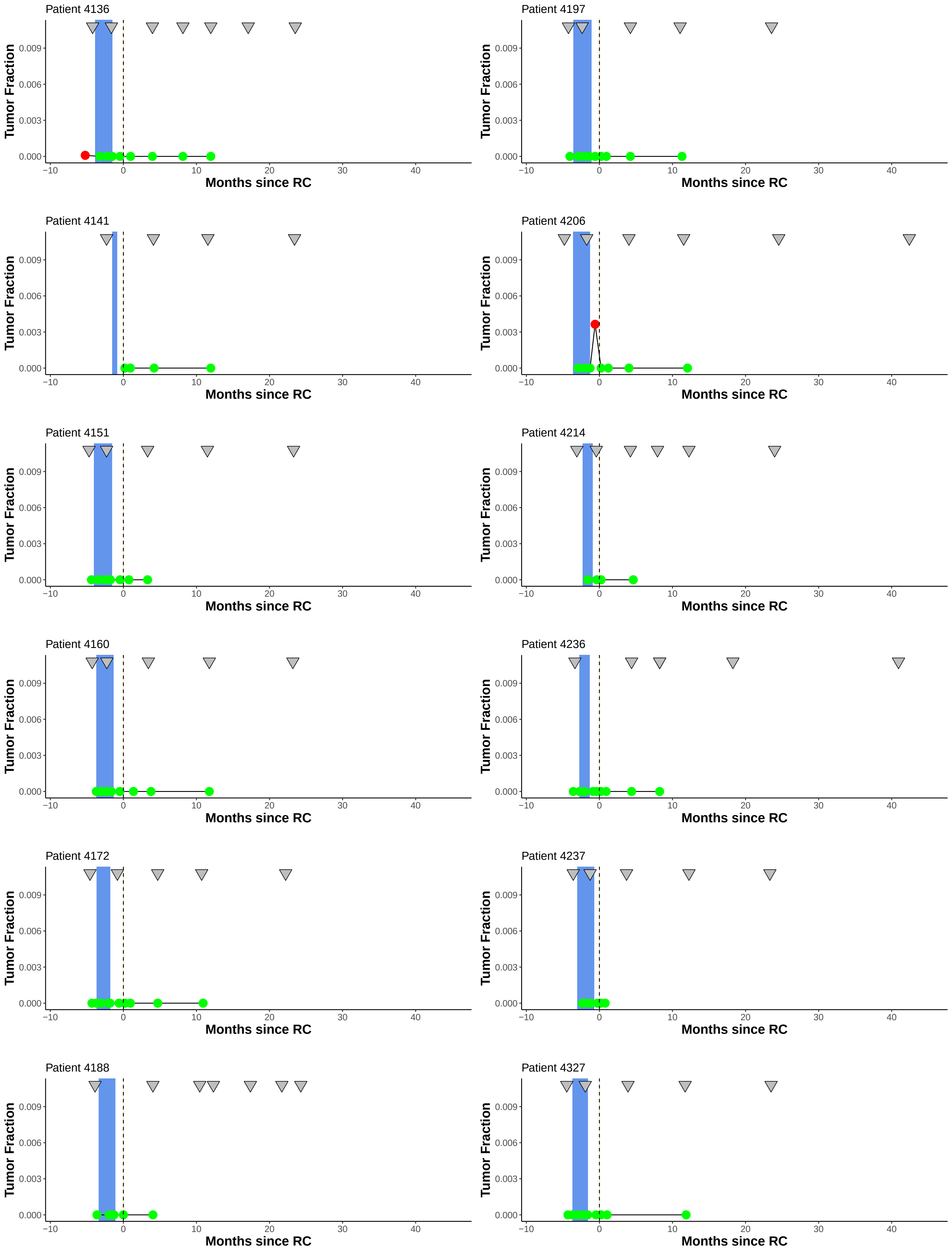
Patient 4136
Patient 4197
Patient 4141
Patient 4206
Patient 4151
Patient 4214
Patient 4160
Patient 4236
Patient 4172
Patient 4237
Patient 4188
Patient 4327
Tumor Fraction
0.000
0.003
0.006
0.009
Months since RC
−10
0
10
20
30
40

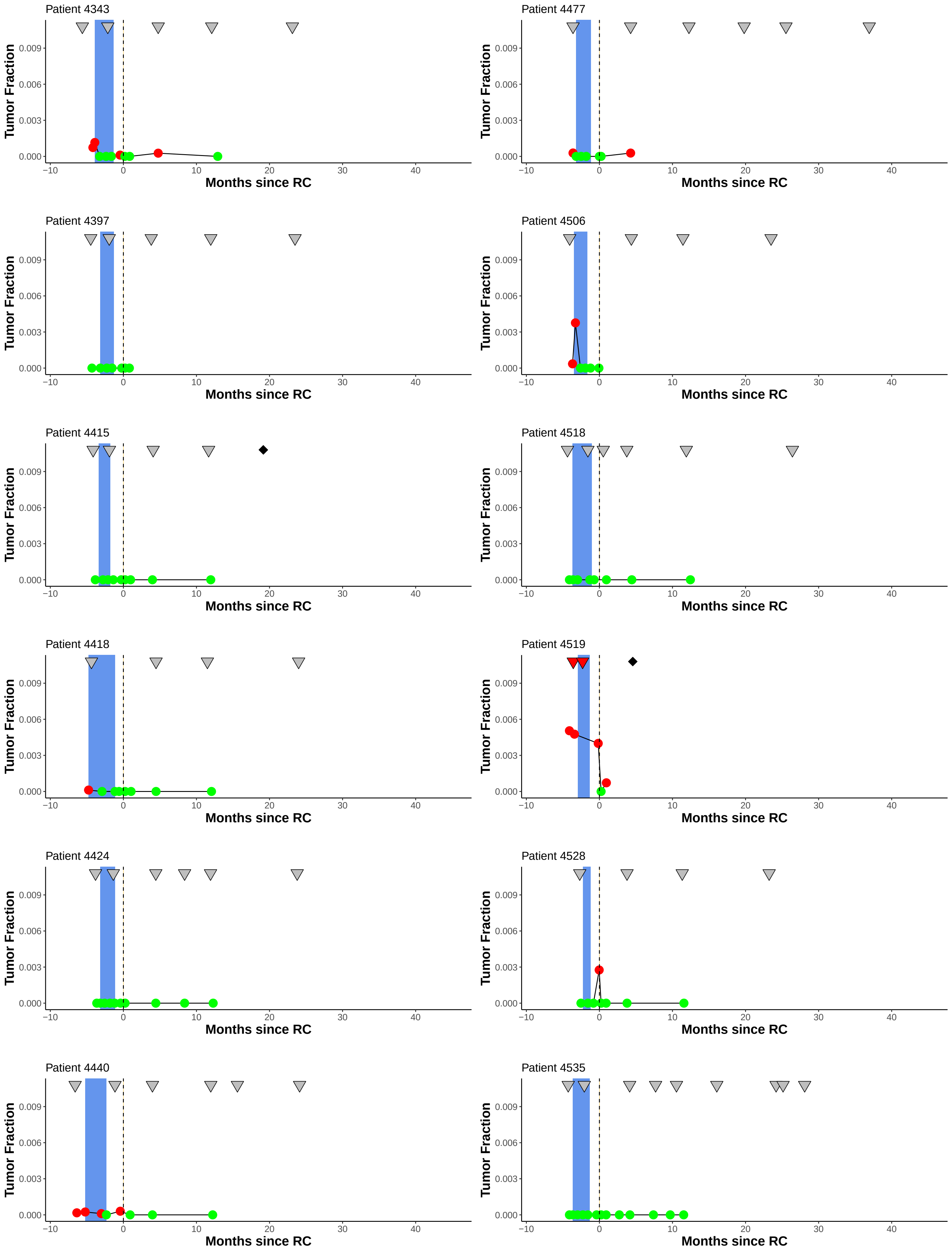
Patient 4343
Tumor Fraction
0.009
0.006
0.003
0.000
−10
0
10
20
30
40
Months since RC
Patient 4477
Tumor Fraction
Months since RC
Patient 4397
Tumor Fraction
Months since RC
Patient 4506
Tumor Fraction
Months since RC
Patient 4415
Tumor Fraction
Months since RC
Patient 4518
Tumor Fraction
Months since RC
Patient 4418
Tumor Fraction
Months since RC
Patient 4519
Tumor Fraction
Months since RC
Patient 4424
Tumor Fraction
Months since RC
Patient 4528
Tumor Fraction
Months since RC
Patient 4440
Tumor Fraction
Months since RC
Patient 4535
Tumor Fraction
Months since RC

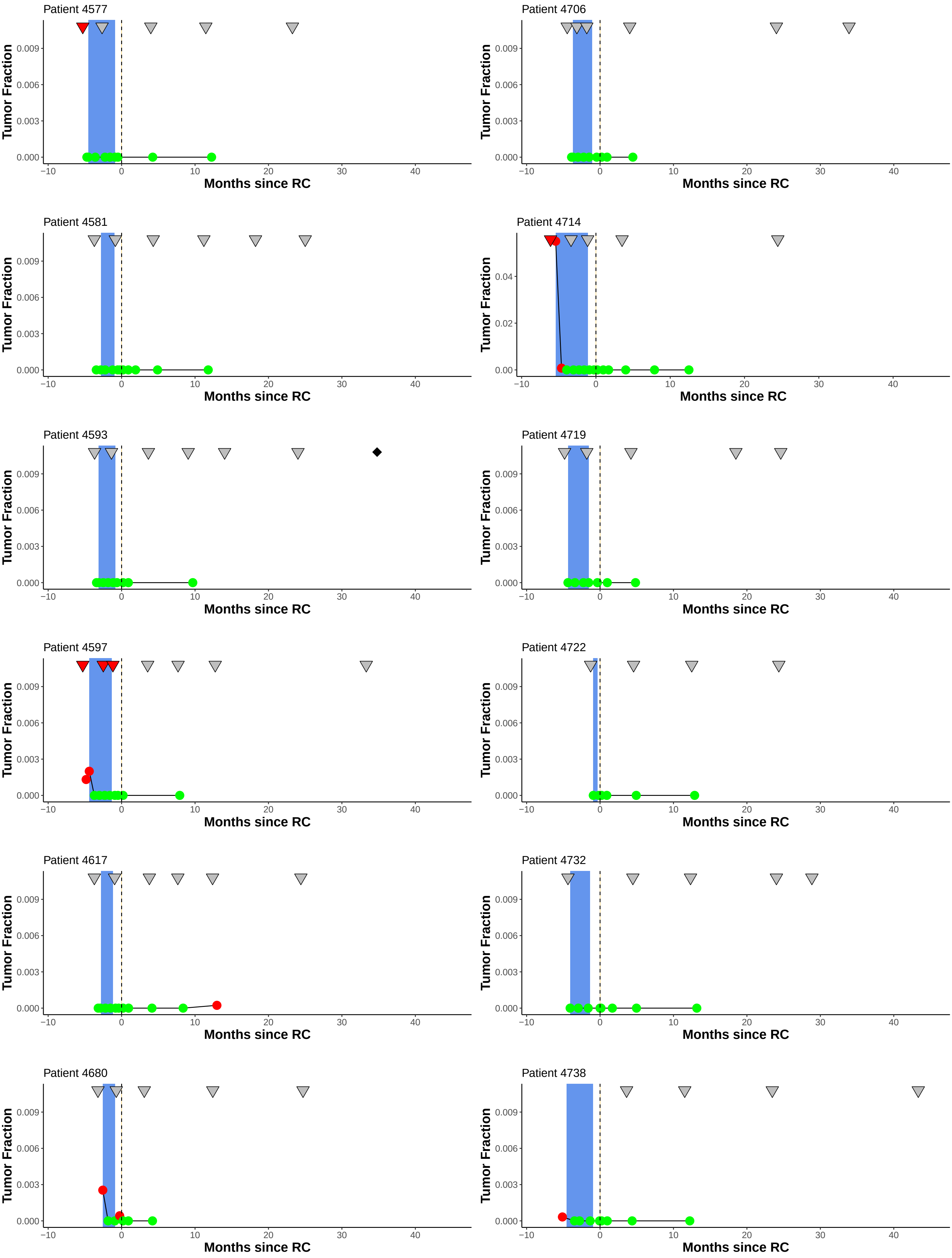

Patient 4577
Patient 4706
Patient 4581
Patient 4714
Patient 4593
Patient 4719
Patient 4597
Patient 4722
Patient 4617
Patient 4732
Patient 4680
Patient 4738
Tumor Fraction
Months since RC
0.000
0.003
0.006
0.009
0.00
0.02
0.04
−10
0
10
20
30
40

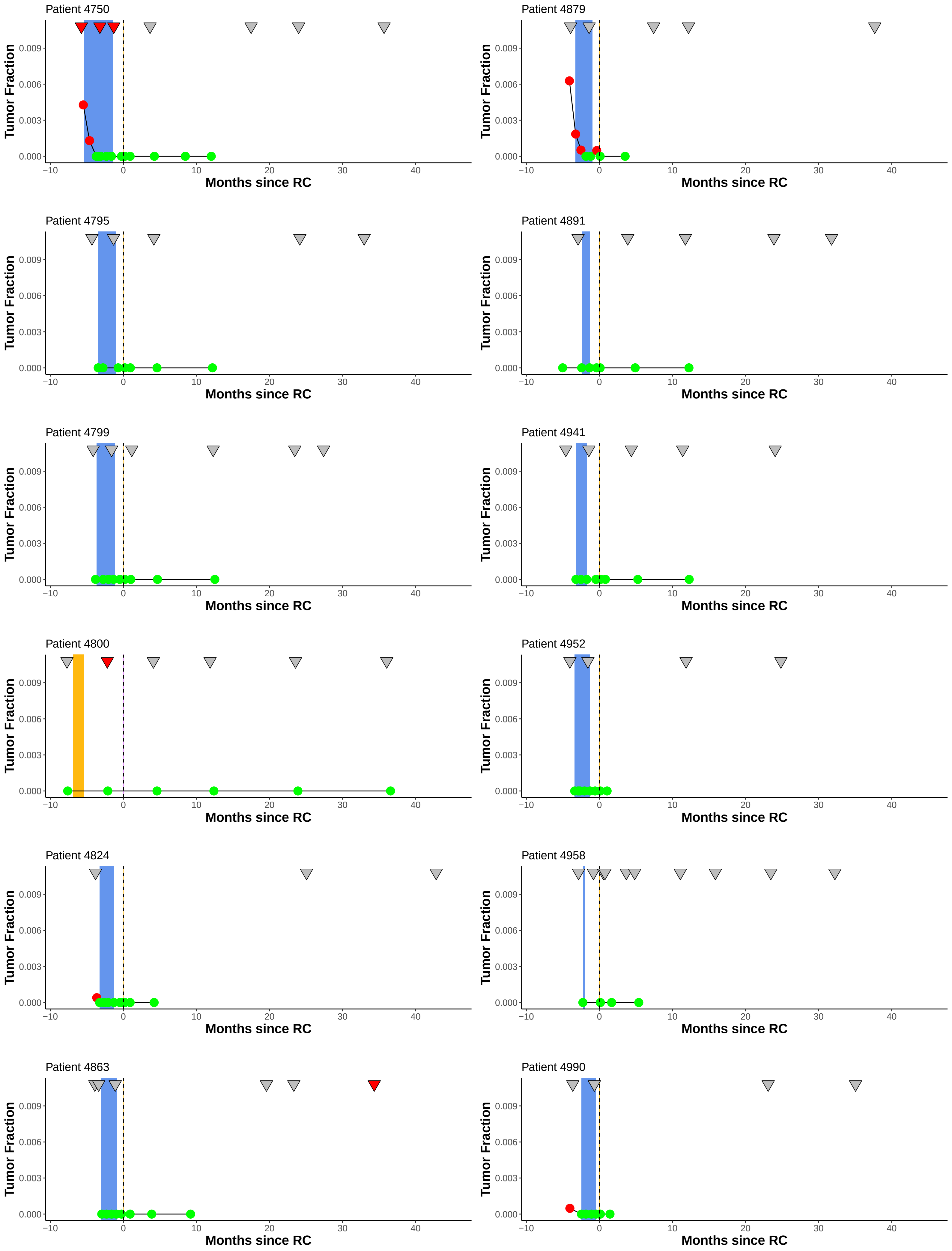
Patient 4750
Patient 4879
Patient 4795
Patient 4891
Patient 4799
Patient 4941
Patient 4800
Patient 4952
Patient 4824
Patient 4958
Patient 4863
Patient 4990
Tumor Fraction
0.000
0.003
0.006
0.009
Months since RC
−10
0
10
20
30
40

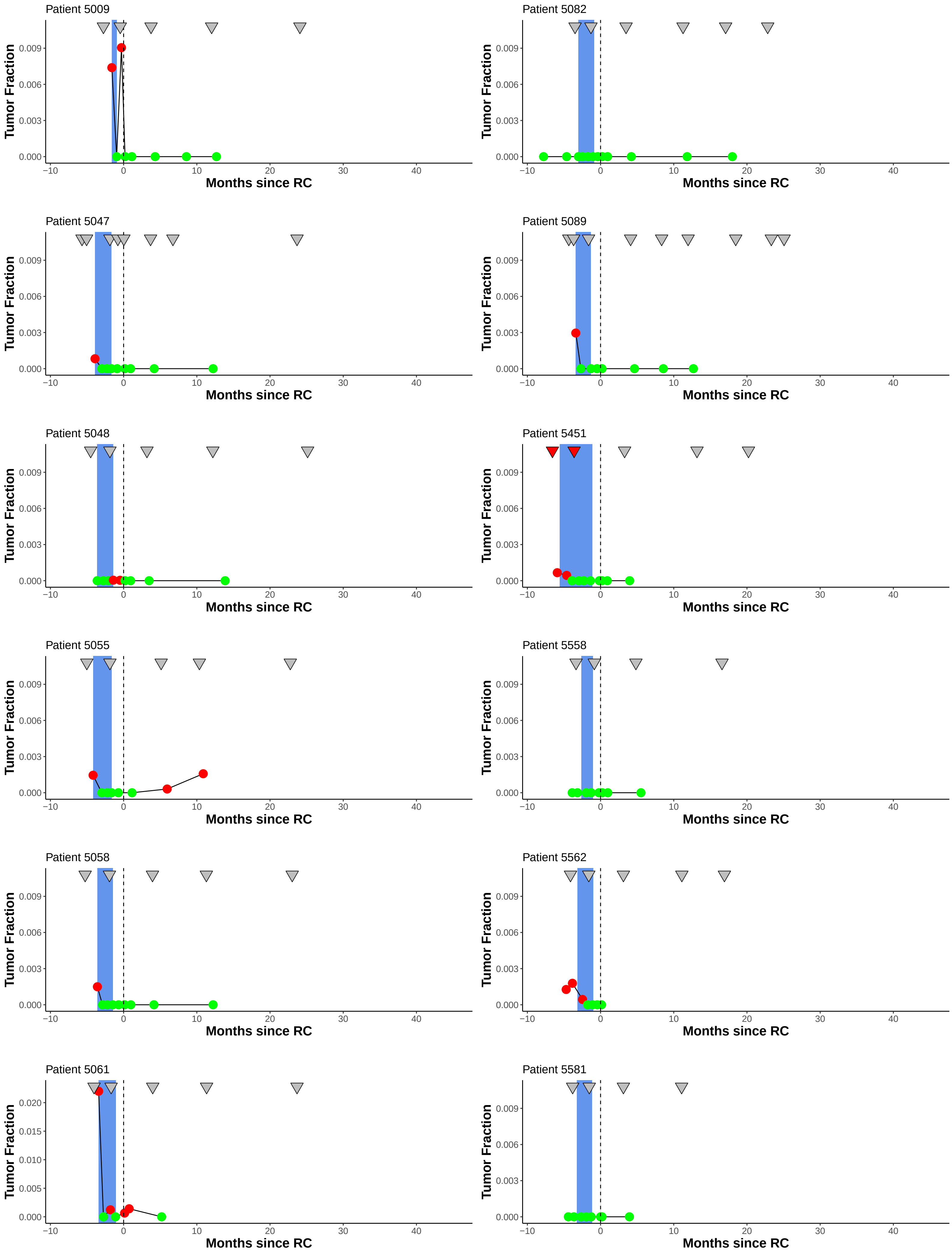

Patient 5009
Patient 5082
Patient 5047
Patient 5089
Patient 5048
Patient 5451
Patient 5055
Patient 5558
Patient 5058
Patient 5562
Patient 5061
Patient 5581
Tumor Fraction
Months since RC
0.000
0.003
0.006
0.009
0.005
0.010
0.015
0.020
−10
0
10
20
30
40

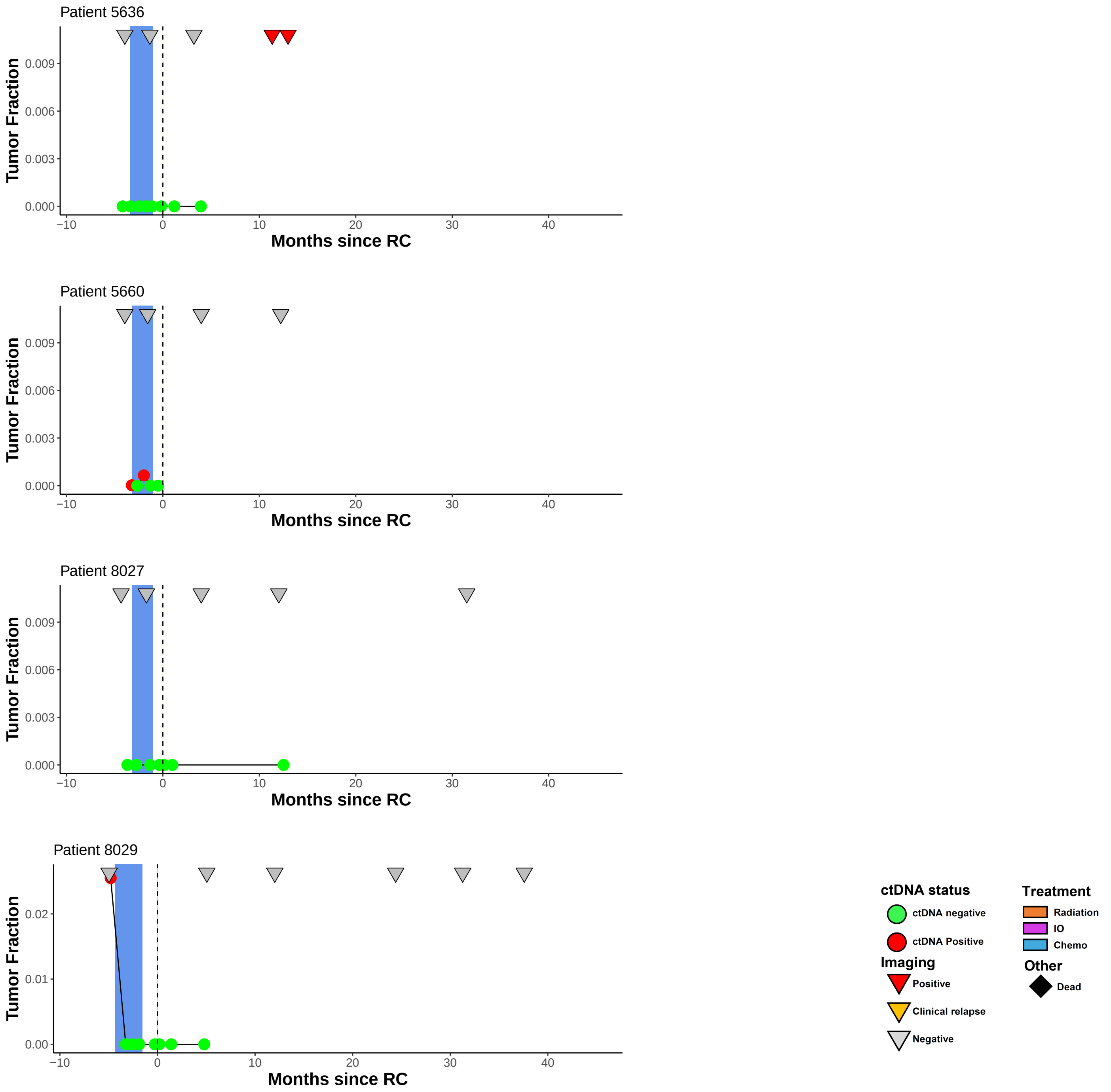


**Supplementary Figure S1**: Detailed disease courses and ctDNA analysis results for all patients. Detailed disease courses and ctDNA Tumor fraction analysis results for all patients. Representation of plasma samples, treatment regimens and imaging results for all patients. The date of planned cystectomy was used for patients 4175 and 4250, however the procedure was not completed. RC = radical cystectomy.

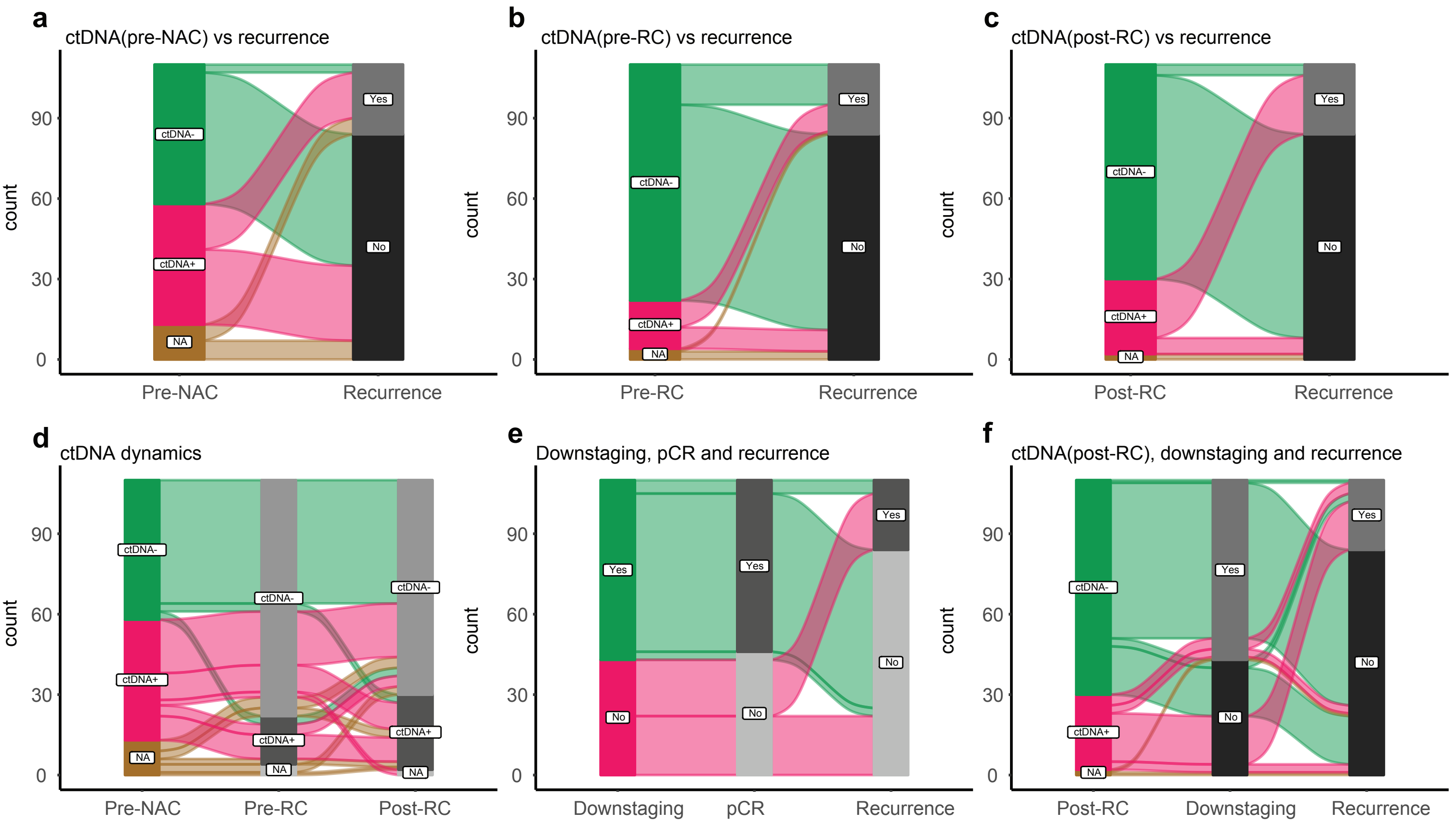


**Supplementary Figure S2. Correlations between ctDNA, pathologic response and recurrence.**
**a-c**, Alluvial plots showing the association between clinical recurrence and ctDNA status at pre-NAC, pre-RC and post-RC. **d,** Alluvial plot showing ctDNA dynamics during the patients' disease courses at the pre-NAC, pre-RC and post-RC time points. **e,** Alluvial plot showing association between pathologic downstaging, pathologic complete response (pCR) and clinical recurrence. **f**, Alluvial plot of association between post-RC ctDNA status, downstaging and clinical recurrence.

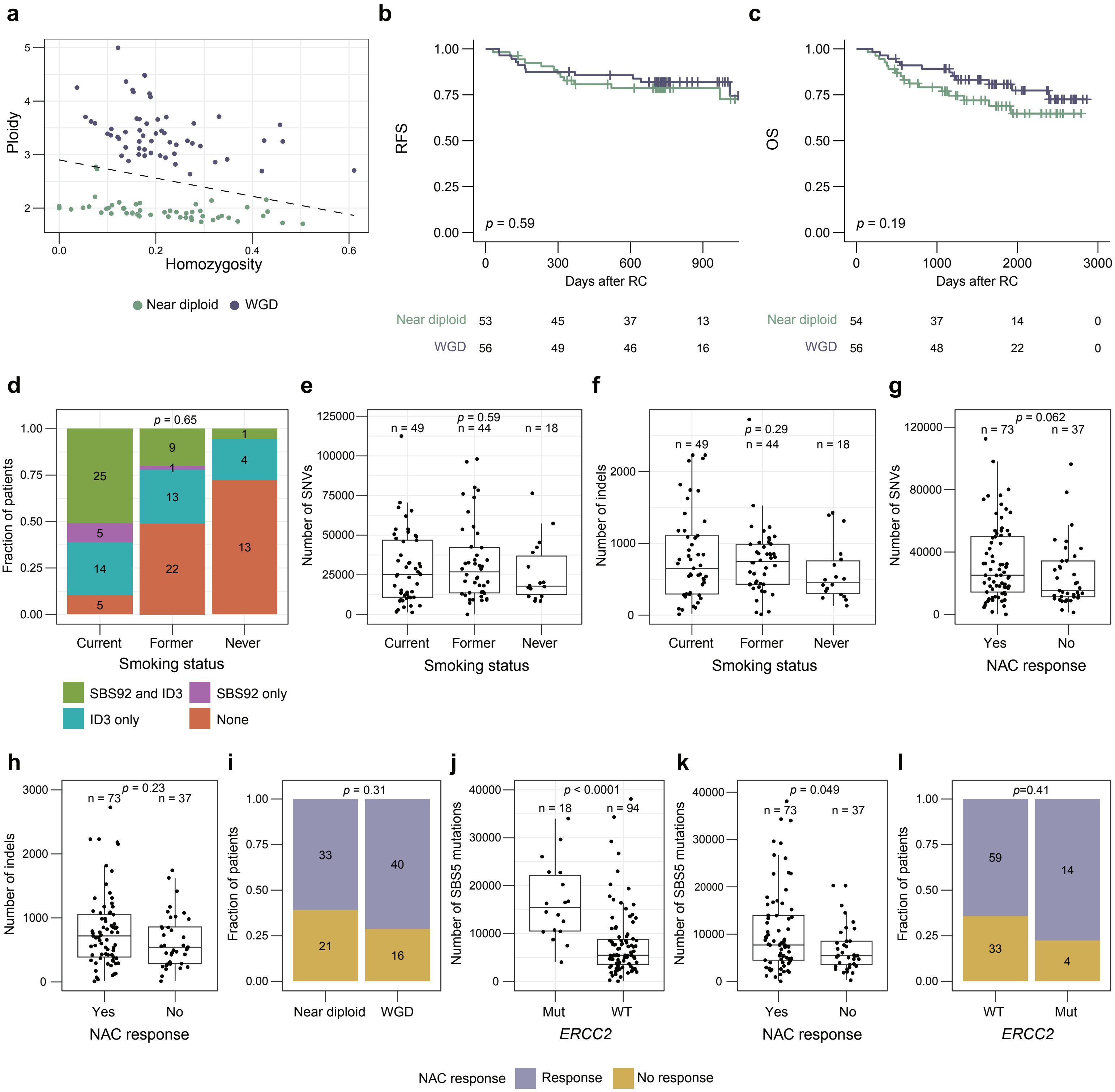

**Supplementary Figure S3: Genomic characterization of primary tumors.**
**a,** Classification of the whole-genome doubling (WGD) status of tumors using ploidy and level of homozygosity. **b,** Kaplan-Meier survival analysis of recurrence-free survival (RFS) of patients stratified by WGD status. **c,** Kaplan-Meier survival analysis of overall survival (OS) of patients stratified by WGD status. **d,** Association between the number of mutations in the SBS92 context and number of small insertions and deletions (indels) in the ID3 context. **e,** Number of single-nucleotide variants (SNVs) according to smoking status of the patients. **f,** Number of indels according to smoking status of the patients. **g,** Number of SNVs according to pathologic response to neoadjuvant chemotherapy (NAC). **h,** Number of indels according to pathologic response to NAC. **i,** Association between pathologic response to NAC and WGD status. **j,** Number of mutations in a SBS5 context according to *ERCC2* mutational status. **k,** Number of mutations in a SBS5 context according to pathologic response to NAC. **l,** Association between pathologic response to NAC and *ERCC2* mutational status.

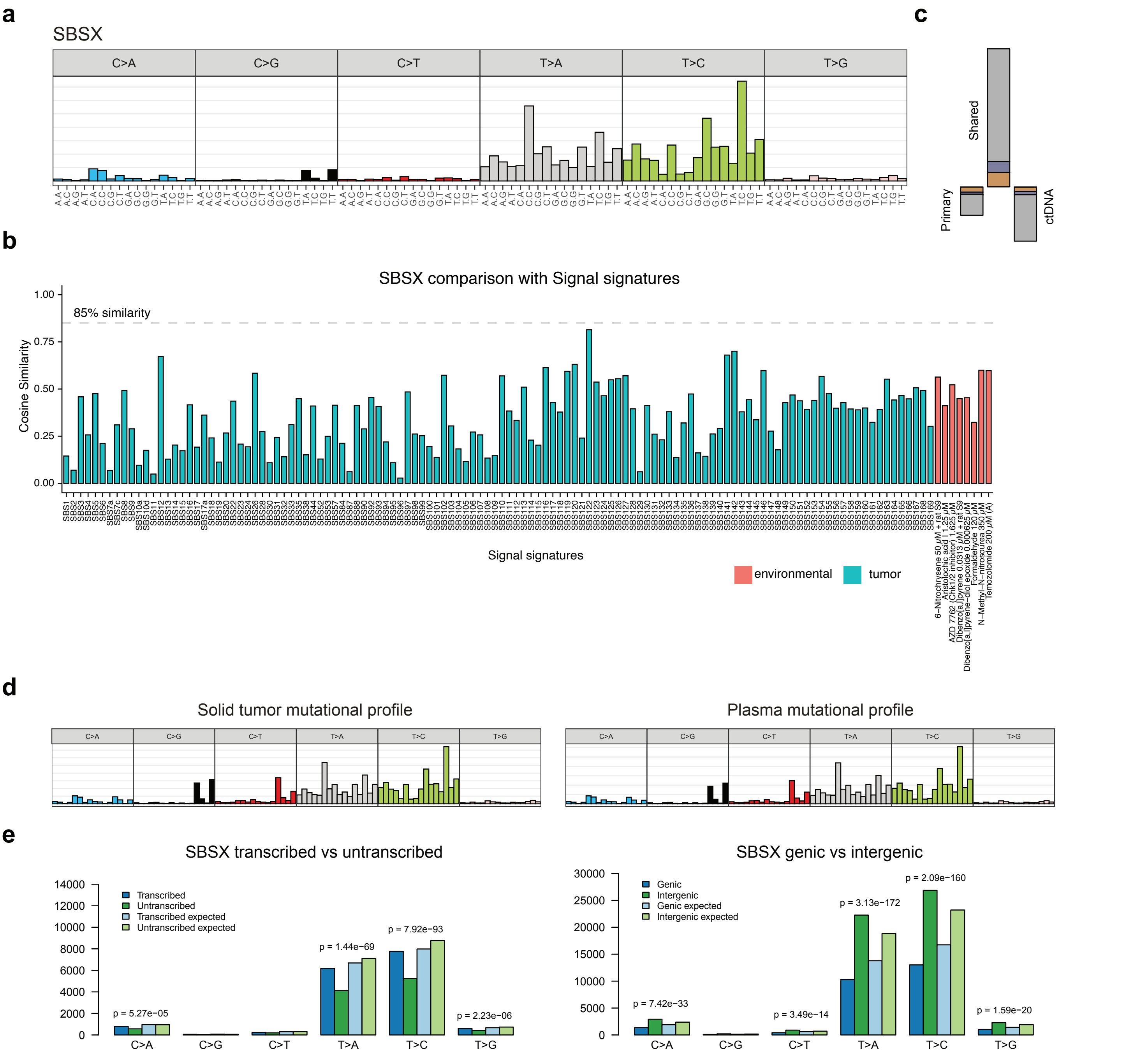


**Supplementary Figure S4: Characterization of a novel SBSX signature.**
**a,** Mutational profile using the conventional 96 mutation type classification for the new signature (SBSX) obtained from sigprofiler. **b,** Cosine similarity between the SBSX signature and Signal signatures. **c,** Contribution of SBSX in primary tumor exclusive mutations (left bar), ctDNA exclusive mutations (right bar), and shared mutations (middle bar). **d,** Mutational profile using the conventional 96 mutation type classification for mutations of the primary tumor sample and the plasma ctDNA. **e,** Enrichment of SBSX activity in transcribed and intergenic regions using sigprofiler topography.

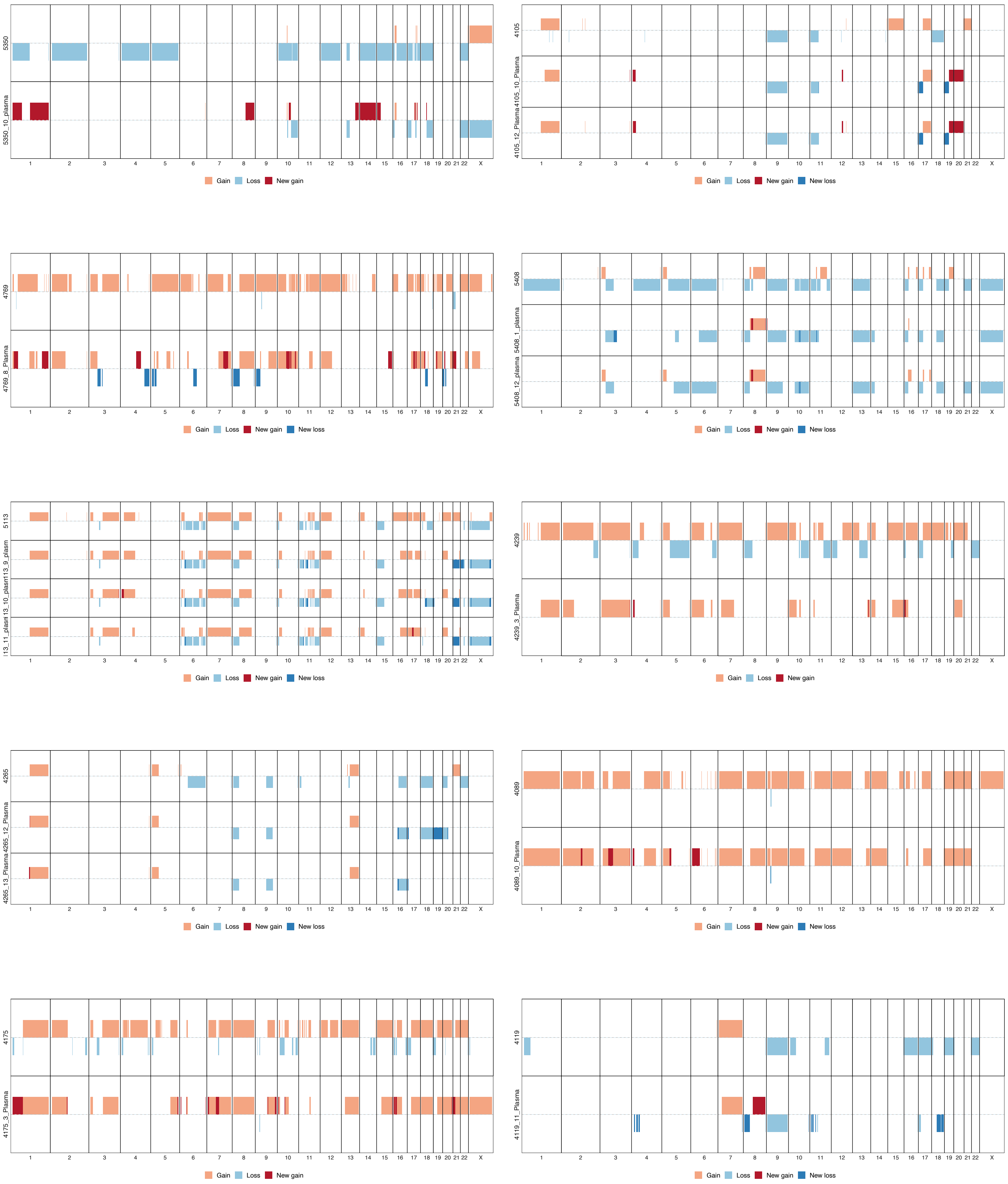


**Supplementary Figure S5: Evolution of copy number changes in plasma.**
Changes in copy numbers comparing primary (top row) versus ctDNA (following rows) for patients having plasma samples with a tumor fraction >10%.

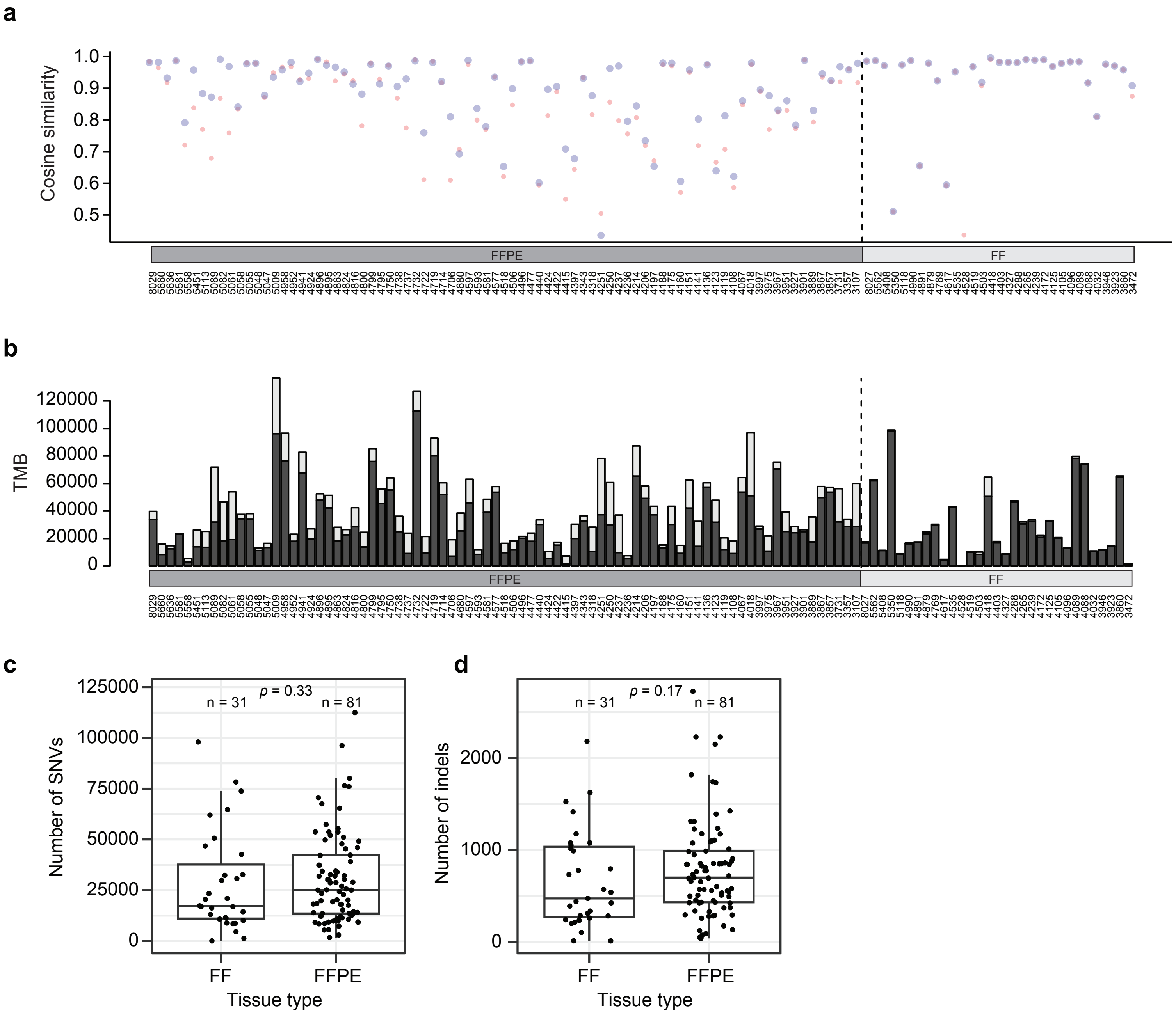


**Supplementary Figure S6: Mutational context similarity and TMB before and after removal of FFPE-induced artifacts.** **a,** Mutational trinucleotide cosine similarity of samples compared with the mean trinucleotide context of fresh frozen (FF) samples before (red dot) and after (blue dot) removal of FFPE-induced artifacts. The median cosine similarity for FFPE samples were 0.87 and 0.92 before and after removal of FFPE-induced artifacts, respectively. **b,** Comparison of tumor mutational burden (TMB) for each sample before (light grey bar) and after (dark grey bar) removal of FFPE-induced artifacts. The median percentage of removed mutations were 29% for FFPE samples compared to 2.7% for FF samples. **c,** Number of SNVs for FF and FFPE samples removal of FFPE-induced artifacts. **d,** Number of indels for FF and FFPE samples after removal of FFPE-induced artifacts.

### Supplementary Table S1. Patient Characteristics and Demographics (n=112)

**Patients, n = 112, median age = 67.0 years (range 42-82)**

| | | | |
|---|---|---|---|
| **Sex, n (%)** | | **smoking status** | |
| Male | 88 (77.9) | current | 49 (43.4) |
| Female | 24 (21.2) | former | 45 (38.8) |
| | | never | 18 (15.9) |
| **pre-therapeutic staging at TUR-B, n (%)** | | **post-therapeutic staging at RC, n (%) (n=110)** | |
| pT1 | 8 (7.1) | ypT0/ypTIS/ypTa | 67 (60.4) |
| pT2 | 95 (84.1) | ypT1 | 7 (6.3) |
| pT3 | 1 (0.9) | ypT2 | 13 (11.7) |
| pT4a | 4 (3.5) | ypT3 | 17 (15.2) |
| pT4b | 3 (2.7) | ypT4a | 6 (5.4) |
| | | ypT4b | 1 (0.9) |
| **pre-therapeutic N stage, n (%)(n=112)** | | **post-therapeutic N staging at RC, n (%) (n=110)** | |
| N0 | 98 (87.5) | ypN0 | 98 (89.1) |
| N1 | 11 (9.8) | ypN1 | 4 (3.6) |
| N2 | 3 (2.7) | ypN2 | 5 (4.5) |
| | | ypN3 | 3 (2.7) |
| **chemotherapy before RC** | | **Median Clinical follow-up, days (range)** | |
| neoadjuvant chemotherapy (NAC) | 112 (100) | Disease free (RC; n=84) | 1841 (139-2810) |
| pathologic downstaging (≤Ta,CIS,N0) | 67 (60.4) | Clinical relapse (RC; n=26) | 705 (198-2493) |
| no pathologic downstaging (≥T1 and/or ≥N1) | 44 (39.6) | Progression (RC impossible; n=2, no post RC imaging; n=1) | |

Supplementary Table S2: Patient characteristics, clinicopathological parameters and ctDNA status stratified for recurrence.

| | Patient characterictics | | | Univariate | | | | | Multivariable/PreNAC, n=88 | | | | Multivariable/PreRC, n=97 | | | | Multivariable/PostRC, n=100 | | | |
|---|---|---|---|---|---|---|---|---|---|---|---|---|---|---|---|---|---|---|---|---|
| **Variable** | **N** | Non-recurrence, N = 83[1] | Recurrence, N = 26[1] | **N** | **HR**[2] | **95% CI**[2] | **p-value** | **q-value**[3] | **N** | **HR**[2] | **95% CI**[2] | **p-value** | **N** | **HR**[2] | **95% CI**[2] | **p-value** | **N** | **HR**[2] | **95% CI**[2] | **p-value** |
| **Age** | 109 | | | | | | | | | | | | | | | | | | | |
| <70 | | 48 (58%) | 19 (73%) | 67 | — | — | | | | | | | | | | | | | | |
| ≥70 | | 35 (42%) | 7 (27%) | 42 | 0.57 | 0.24, 1.35 | 0.2 | 0.2 | | | | | | | | | | | | |
| **Sex** | 109 | | | | | | | | | | | | | | | | | | | |
| F | | 16 (19%) | 8 (31%) | 24 | — | — | | | | | | | | | | | | | | |
| M | | 67 (81%) | 18 (69%) | 85 | 0.57 | 0.25, 1.32 | 0.2 | 0.2 | | | | | | | | | | | | |
| **Smoking** | 109 | | | | | | | | | | | | | | | | | | | |
| Never | | 11 (13%) | 6 (23%) | 17 | — | — | | | | | | | | | | | | | | |
| Former | | 34 (41%) | 10 (38%) | 44 | 0.54 | 0.20, 1.49 | 0.2 | 0.3 | | | | | | | | | | | | |
| Current | | 38 (46%) | 10 (38%) | 48 | 0.47 | 0.17, 1.30 | 0.15 | 0.2 | | | | | | | | | | | | |
| **Downstaging** | 109 | | | | | | | | | | | | | | | | | | | |
| Response | | 67 (81%) | 6 (23%) | 73 | — | — | | | 59 | — | — | | 65 | — | — | | 69 | — | — | |
| No response | | 16 (19%) | 20 (77%) | 36 | 12.1 | 4.78, 30.5 | **<0.001** | <0.001 | 30 | 10.0 | 2.56, 39.2 | **<0.001** | 33 | 11.7 | 4.26, 32.2 | **<0.001** | 32 | 11.9 | 3.40, 42.0 | **<0.001** |
| **ypT(RC)** | 109 | | | | | | | | | | | | | | | | | | | |
| T0-Ta | | 55 (66%) | 5 (19%) | 60 | — | — | | | | | | | | | | | | | | |
| T1-T4 | | 28 (34%) | 21 (81%) | 49 | 9.26 | 3.42, 25.1 | **<0.001** | <0.001 | | | | | | | | | | | | |
| **ypN(RC)** | 108 | | | | | | | | | | | | | | | | | | | |
| N0 | | 81 (98%) | 16 (64%) | 97 | — | — | | | | | | | | | | | | | | |
| N1-N3 | | 2 (2%) | 9 (36%) | 11 | 22.1 | 8.51, 57.5 | **<0.001** | <0.001 | | | | | | | | | | | | |
| **PreNAC ctDNA** | 96 | | | | | | | | | | | | | | | | | | | |
| Negative | | 49 (64%) | 3 (15%) | 52 | — | — | | | 48 | — | — | | | | | | | | | |
| Positive | | 27 (36%) | 17 (85%) | 44 | 9.18 | 2.68, 31.5 | **<0.001** | 0.001 | 41 | 2.91 | 0.74, 11.4 | 0.13 | | | | | | | | |
| **PreRC ctDNA** | 105 | | | | | | | | | | | | | | | | | | | |
| Negative | | 72 (90%) | 15 (60%) | 87 | — | — | | | | | | | 80 | — | — | | | | | |
| Positive | | 8 (10%) | 10 (40%) | 18 | 3.86 | 1.73, 8.61 | **<0.001** | 0.002 | | | | | 18 | 2.25 | 0.98, 5.14 | 0.055 | | | | |
| **PostRC ctDNA** | 107 | | | | | | | | | | | | | | | | | | | |
| Negative | | 76 (94%) | 4 (15%) | 80 | — | — | | | | | | | | | | | 74 | — | — | |
| Positive | | 5 (6%) | 22 (85%) | 27 | 31.6 | 10.8, 92.9 | **<0.001** | <0.001 | | | | | | | | | 27 | 45.0 | 9.54, 212 | **<0.001** |
| **ctDNA dynamic** | 38 | | | | | | | | | | | | | | | | | | | |
| Remains | | 3 (12%) | 6 (46%) | 9 | — | — | | | | | | | | | | | | | | |
| Clearance | | 22 (88%) | 7 (54%) | 29 | 0.32 | 0.11, 0.95 | **0.040** | 0.075 | | | | | | | | | | | | |
| **BMI** | 108 | | | | | | | | | | | | | | | | | | | |
| <30 | | 62 (76%) | 17 (65%) | 79 | — | — | | | | | | | | | | | | | | |
| ≥30 | | 20 (24%) | 9 (35%) | 29 | 1.42 | 0.63, 3.18 | 0.4 | 0.4 | | | | | | | | | | | | |
| **NAC cycles** | 109 | | | | | | | | | | | | | | | | | | | |
| <3 | | 11 (13%) | 7 (27%) | 18 | — | — | | | | | | | | | | | | | | |
| ≥3 | | 72 (87%) | 19 (73%) | 91 | 0.55 | 0.23, 1.31 | 0.2 | 0.2 | | | | | | | | | | | | |
| **RFS(days)** | 109 | 1,852 (1,253, 2,398) | 706 (395, 1,298) | | | | | | | | | | | | | | | | | |

[1] n (%); Median (IQR)

[2] HR = Hazard Ratio, CI = Confidence Interval

[3] False discovery rate correction for multiple testing

Patient characteristics for patients compleated RC (patient 4519 excluded due to death before first post RC imaging).
Univariate and multivariable analysis were stratified for recurrence.

### Supplementary Table S3: dndscv results

| | Alterations (n) | | | | | |
|---|---|---|---|---|---|---|
| Gene | Synonomous | Missense | Nonsense | Splice site | Indels | Global q-value |
| *TP53* | 3 | 60 | 6 | 3 | 10 | 0 |
| *RB1* | 0 | 5 | 11 | 11 | 12 | 0 |
| *KDM6A* | 2 | 4 | 15 | 3 | 5 | 0 |
| *ARID1A* | 3 | 9 | 13 | 1 | 15 | 0 |
| *ELF3* | 1 | 6 | 2 | 0 | 10 | 1.28E-12 |
| *CDKN1A* | 0 | 1 | 2 | 0 | 8 | 1.17E-11 |
| *FOXQ1* | 0 | 2 | 1 | 0 | 10 | 3.87E-10 |
| *CDKN2A.p16INK4a* | 0 | 1 | 3 | 0 | 2 | 0.000112568 |
| *ERCC2* | 1 | 18 | 0 | 0 | 0 | 0.000516769 |
| *PIK3CA* | 1 | 22 | 0 | 0 | 0 | 0.000516769 |
| *CDKN1B* | 2 | 1 | 2 | 0 | 3 | 0.001424424 |
| *KMT2D* | 10 | 16 | 11 | 1 | 5 | 0.00146858 |
| *FGFR3* | 1 | 14 | 0 | 0 | 1 | 0.009272726 |
| *ERBB2* | 2 | 20 | 0 | 1 | 0 | 0.013244694 |
| *TSC1* | 0 | 1 | 3 | 0 | 4 | 0.014328416 |
| *ACTB* | 0 | 9 | 0 | 0 | 1 | 0.014328416 |
| *KLF5* | 1 | 6 | 1 | 0 | 2 | 0.021455963 |
